# Not all controls are made equal: Definition of human kidney reference samples by single cell gene expression profiles

**DOI:** 10.1101/2025.03.17.25324134

**Authors:** Rajasree Menon, Paul L. Kimmel, Edgar A. Otto, Lalita Subramanian, Christopher L. O’ Connor, Bradley Godfrey, Cathy Smith, Fadhl Alakwaa, Celine C. Berthier, Minnie Sarwal, E. Steve Woodle, Laura Pyle, Ye Ji Choi, Patricia Ladd, John R. Sedor, Syvia E. Rosas, Sushrut S. Waikar, Abhijit S. Naik, Ricardo Melo Ferreira, Michael T. Eadon, Markus Bitzer, Petter Bjornstad, Jeffrey B. Hodgin, Matthias Kretzler, Kidney Precision Medicine Project (KPMP)

## Abstract

Identifying mechanisms of kidney disease typically involves comparing diseased samples to healthy reference tissues. However, the impact of variability in tissue procurement, storage, and donor characteristics remains incompletely explored. In this study, we evaluated three kidney reference types: tumor nephrectomy (TN), pre-transplant biopsies from living donor (LD), and percutaneous biopsies from healthy control volunteers (HC) for their influence on differential gene expression across three diabetic kidney disease (DKD) states. We identified distinct injury markers, cell state proportions, and gene signatures linked to procurement procedures and donor age. Adjusting for these confounders altered pathway analysis results. For example, fatty acid metabolism was most enriched in HC samples after age correction, while an interferon gamma response in the diabetes mellitus resilient (DM-R; patients with diabetes and minimal kidney impairment) vs. HC comparison disappeared. Biological processes associated with aging were enriched in older reference tissues, potentially overlapping with disease mechanisms. Notably, tumor necrosis factor signaling via nuclear factor-κB remained enriched in LD and TN compared to HC, even after adjustment for age and procurement. These findings emphasize the importance of selecting appropriate control tissues to accurately identify kidney disease mechanisms.

## INTRODUCTION

The heterogeneous nature of kidney diseases has hindered our understanding of disease pathogenesis. Advances in genomics technologies have begun to unveil the molecular mechanisms underlying kidney diseases (1–4), yet these discoveries relied heavily on comparisons between diseased and reference tissue. Given the challenges of obtaining disease-free kidney tissue from healthy humans, a wide variety of sources for reference tissue have been used. Moreover, factors such as the sample procurement, preservation method, tissue processing technology, and the age and underlying physiological condition of the donor may substantially impact reference tissue and corresponding gene expression profiles (5).

As molecular analysis tools continue to advance, an understanding of the advantages and limitations of different reference tissue sources has become critical to define the context of comparative analyses. In this study, we compared single-cell gene expression data from three reference tissue sources: percutaneous kidney research biopsies from healthy controls (HC), unaffected parts of tumor-nephrectomies (TN), and perioperative living kidney donor biopsies (LD).

We analyzed single-cell gene expression data from percutaneous kidney research biopsies of patients with type 2 diabetes with diabetic kidney disease (DKD), patients with type 1 diabetes for over 25 years without clinical evidence of kidney disease (hereafter, diabetes mellitus resilient [DM-R]), and youth with early onset type 2 diabetes and kidney disease (early DKD) as disease comparators to determine how reference tissue selection impacts identification of molecular mechanisms driving DKD progression (Figure 1).

**Figure 1.**
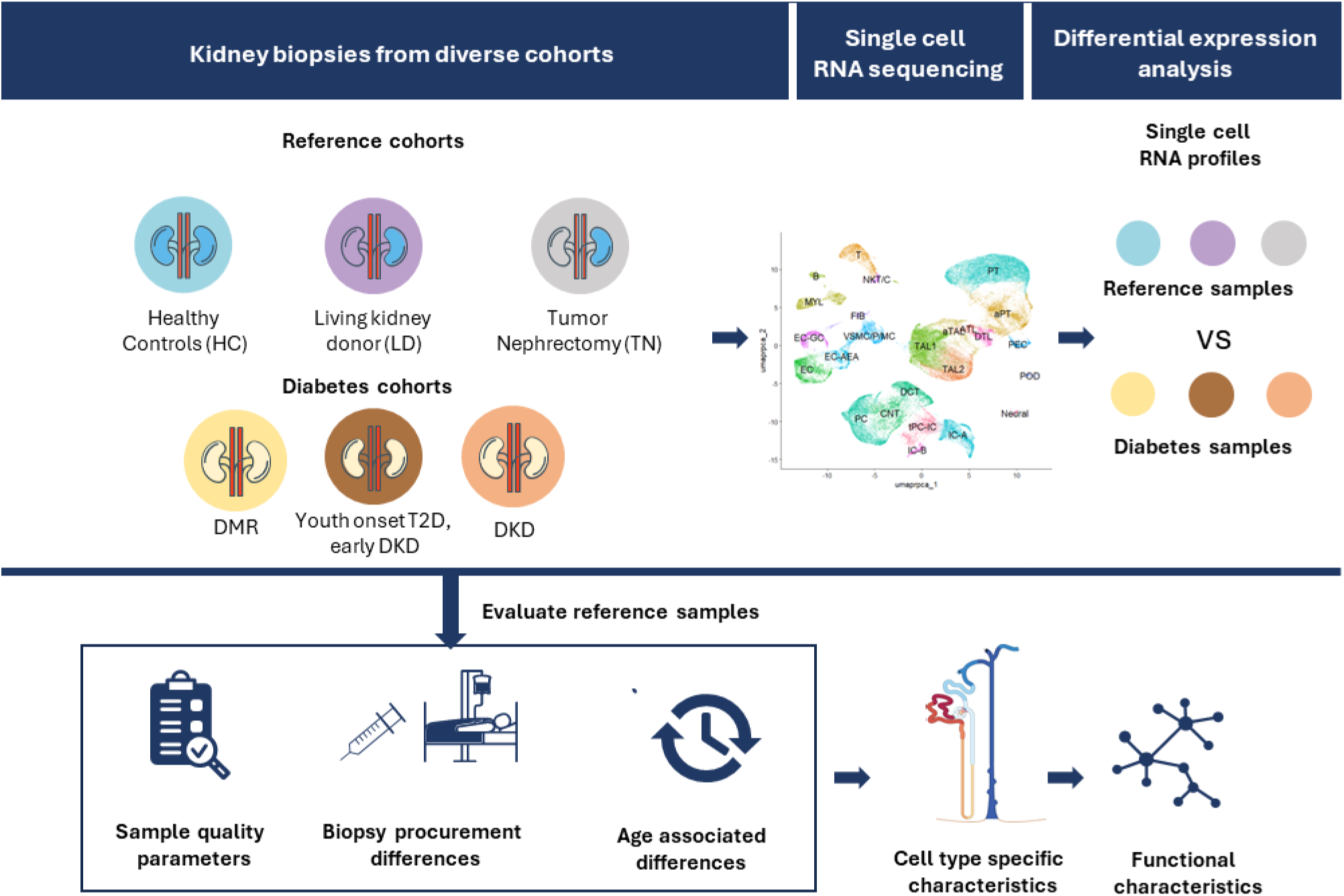
Overview of the analysis approach

## RESULTS

### Single cell and Visium data analysis

In our down-sampled integrated single-cell dataset, comprising 120,000 cells from 74 samples across 6 sample groups, we annotated 23 distinct cell types (Figure 2a). Within these, we identified two cell states for proximal tubule cells, healthy and adaptive/maladaptive (PT and aPT). The aPT state was specifically marked by over-expression of Vascular Cell Adhesion Molecule 1 (VCAM1), a known indicator of adaptive/maladaptive proximal cells (1). For the thick ascending loop of Henle cells (TAL), we identified three states: TAL1, TAL2, and aTAL, with the aTAL state being characterized by high expression of Prominin 1 (PROM1) (1). Importantly, no batch effect was observed in the integrated dataset (Figure 2b, Supplementary Figure 1).

**Figure 2.**
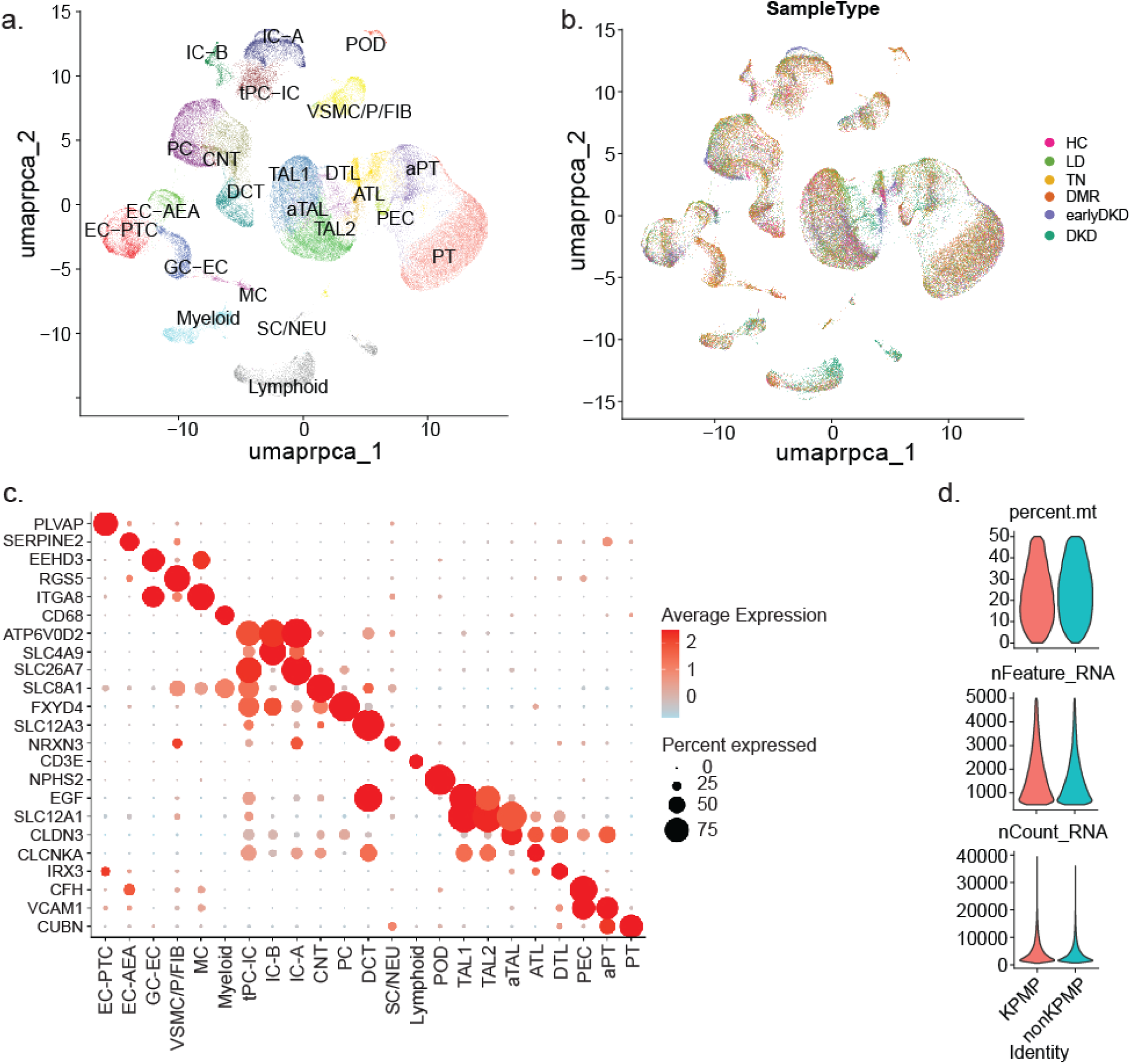
Single cell data. **a.** UMAP showing the annotated cell types from the integrated analysis of a total of 119,193 cells from 6 sample types (20,000 cells each from Living donor “LD”, Tumor-nephrectomy “TN”, Diabetic mellitus resistor “DM-R”, early chronic kidney disease “early DKD”, Diabetic kidney disease “DKD” and 19193 cells from “HC”). Cell type abbreviations include: Podocyte, POD; Parietal epithelial cell, PEC; Proximal tubule, PT; adaptive/maladaptive, aPT; descending thin loop of Henle, DTL; ascending thin loop of Henle, ATL; thick ascending loop of Henle, TAL; adaptive/maladaptive thick ascending loop of Henle, aTAL; distal convoluted tubule, DCT; connecting tubule, CNT; Principal cell, PC; Intercalated type A, IC-A; Intercalated type B, IC-B; transient between PC and IC, tPC-IC; Endothelial cell EC; Efferent and Afferent arteriolar endothelial cells EC-AEA; Glomerular endothelial cell, EC-GC; Fibroblast, FIB; Vascular smooth muscle cell, VSMC; Pericyte, P; Mesangial cell, MC; Myeloid cell, MYL; B cell, B; T cell, T; Natural killer T cell, NK T/C. **b.** UMAP of the integrated dataset showing successful integration of HC, LD, TN, DM-R, early DKD and DKD. **c.** Dot plot showing the specific markers for the annotated cell types. **d**. Violin plots showing the quality control features: percentage of mitochondrial reads per cell (percent.mt), number of features per cell (nFeature_RNA) and number of read counts per cell (nCounts_RNA) in KPMP and non-KPMP samples.

Supplementary Table 1 provides the number of cells per cell type identified from all the samples used in this study. Figure 2c shows the specific expression of markers used to annotate the cell types. Figure 2d shows the violin plots of the generally used single cell quality control (QC) features: Mitochondrial reads percentage per cell, number of genes per cell and number of unique reads per cell for the KPMP and non-KPMP samples, with comparable QC features between KPMP and non-KPMP samples. UMAPs from the integrated analysis of the seventeen 10X-Visium datasets from the 6 sample groups are shown in Supplementary Figure 2. The integrated dataset contained a total of 14,012 spots.

### Early stress and injury/disease markers

First, we assessed tissue stress and injury across the different kidney tissue profiles. The proportion of reads mapping to mitochondrial genes within a cell serves as an essential QC metric and identifies severely stressed and damaged cells (6). Single-cell analysis of kidney tissue shows, compared to other tissues, a high mitochondrial read content. In this study, we included only cells with 50% or fewer reads of the 13-protein coding mitochondrial genes. The dot plot (Figure 3a) summarizes the mitochondrial reads per cell, with a gradient seen from the lowest percentage of reads per cell in HC to TN and LD groups.

**Figure 3.**
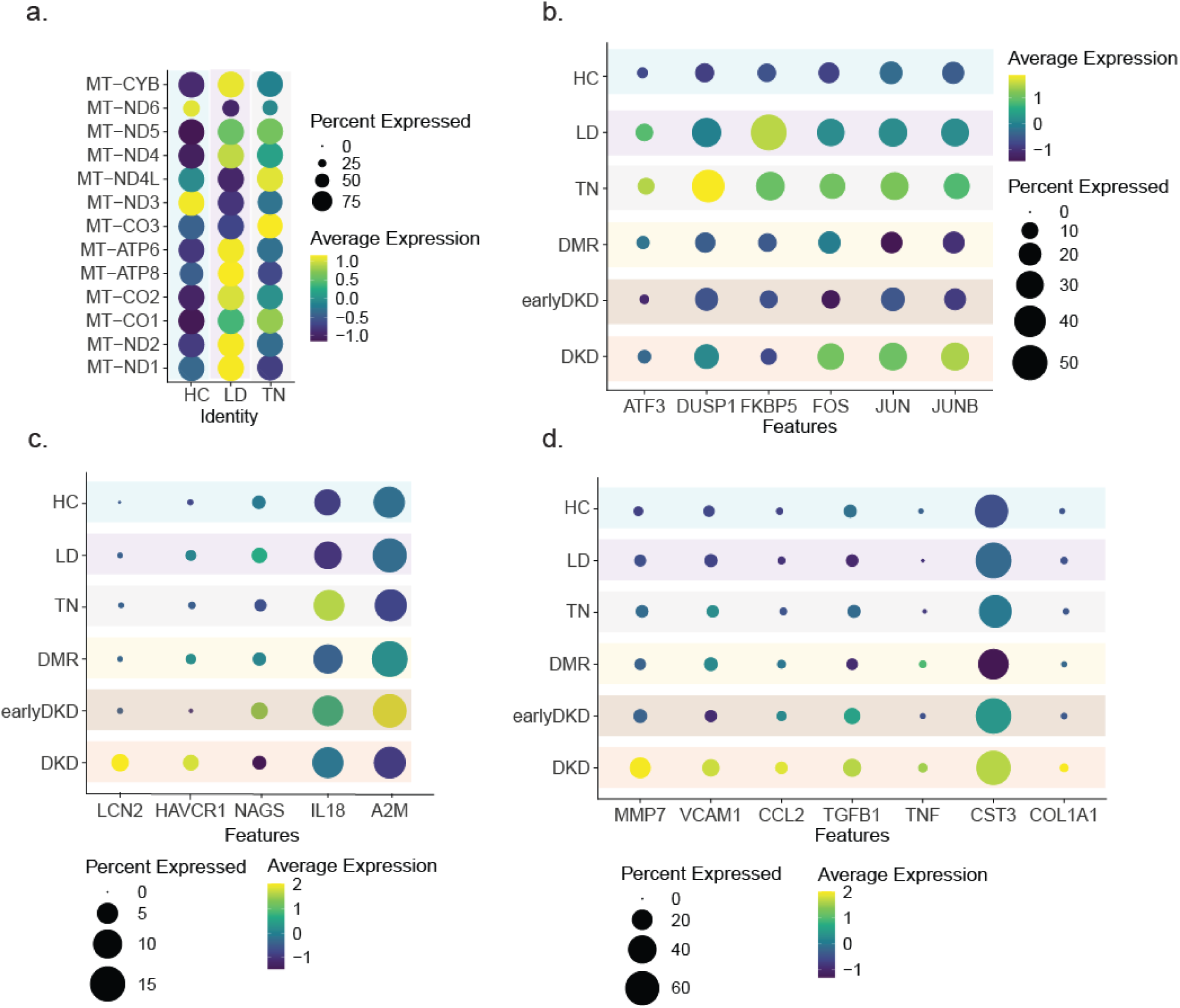
Early stress response and injury // disease markers. **a**. Dot plot depicting relative expression levels of the 13 protein-coding mitochondrial genes across healthy controls (HC), living donors (LD), and tumor nephrectomies (TN). Most mitochondrial genes showed lowest expression in HC and the highest expression in LD. **b**. Dot plot illustrating scaled expression of early stressor genes (ATF3, DUSP1, FOS, JUN, JUNB, FKBP5) across six sample types. Expression was highest in TN and LD. **c**. Dot plot showing expression of early CKD/injury markers in six sample types. Among reference groups, HC exhibited the lowest levels of HAVCR1, LCN2, and IL18. **d**. Dot plot displaying expression of progressive/late disease markers across six sample types. Reference groups had relatively low expression compared to DKD, with little difference observed among reference groups.

Early stress response gene expression can be activated within minutes after stimulation (7, 8) and can serve as a sensitive biomarker of cellular stress. As for Mitochondrial DNA content, a gradient in the early stress response genes was observed. Activating transcription factor 3 (ATF3), Dual Specificity Phosphatase 1 (DUSP1), Early growth response 1 (EGR1), Fos Proto-Oncogen (FOS) and Jun Proto-Oncogene (JUN) were observed lowest in HC and highest in TN among the reference tissues (Figure 3b). Both LD and TN had higher expressions of these genes than DM-R and early DKD.

In addition to the non-specific stress markers studied above, kidney tissue specific markers of cellular stress and damage have been developed and were assessed next. The expression levels of early kidney injury/disease markers including Alpha 2 Macroglobulin (A2M), Hepatitis A virus cellular receptor 1 (HAVCR1, encoding Kidney Injury Marker1 (KIM1)), Interleukin 18 (IL18), Lipocalin-2 (LCN2, encoding NGAL) and N-acetylglutamate synthase (NAGS) are shown in Figure 3c. Among the reference groups, HC had the lowest expression for most of these genes. Although expressed at low levels, HAVCR1 and NAGS markers of kidney damage were detected in LD, compared to the other two reference groups. IL18, an early indicator of acute kidney injury, showed relatively high expression levels in TN. No expression of LCN2, another early AKI marker was observed in any of the reference groups.

Finally, CKD markers of chronic tissue stress and damage were evaluated. No differences in the late CKD markers, including Chemokine (C-C motif) Ligand 2 (CCL2), Collage A1 (COL1A1), Cystatin C (CST3), Matrix Metalloproteinase 7 (MMP7), Transforming Growth Factor Beta 1 (TGFB1), Tumor Necrosis Factor (TNF) and VCAM1, were observed among the reference tissue groups (Figure 3d).

### Cell type proportion

Figure 4a and Supplementary Figure 3 illustrate the proportions of cell types across sample groups. Relative to DKD, the reference groups exhibited substantially lower proportions of aPT, aTAL, lymphoid, and myeloid cells, as demonstrated by the heatmap (Figure 4b) generated from mean cell type proportions. The observed heterogeneity among aPT, aTAL, and immune cells is consistent with findings from recent single-cell studies (1, 9, 10), for example, only very small B cell population was found in HC compared to LD and TN (Figure 4c).

**Figure 4.**
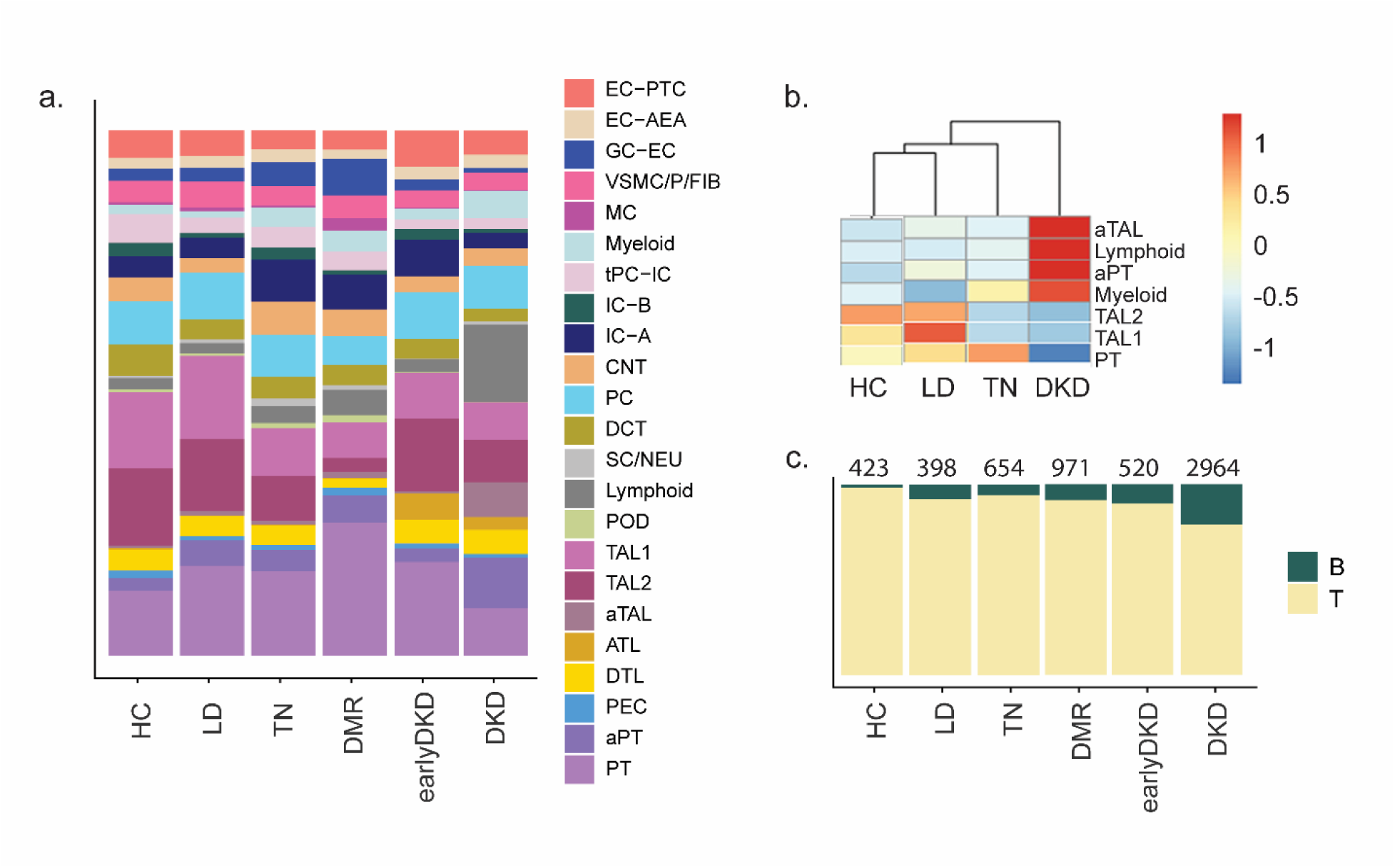
Immune cells and tubular epithelial cell states. **a.** Proportion of cell types found in each of the 6 sample types (HC, LD, TN, DM-R, early DKD and DKD). **b.** The mean proportion of PT, aPT, TAL1, TAL2, aTAL, lymphoid, and myeloid per sample groups studied shown in a heatmap. The scaled expression levels show the most in DKD compared to reference groups. **c.** The proportions of T and B cells within the lymphoid cell population were assessed across six sample groups. B cell proportion was lowest in healthy controls (HC) and highest in diabetic kidney disease (DKD).

### Effect of post-operative tissue procurement

Differential expression analysis between the post-operative procured sample types (LD and TN) to the percutaneous biopsy procurement (HC, all DKD samples), along with manual curation, identified a set of 25 genes (Supplementary Table 2) that exhibited significantly elevated expression in the samples from post-operative surgical reference tissue (Figure 5a,5b). Network analysis using STRING (11) revealed direct interactions among 19 of these 25 genes (Figure 5c), with enrichment in the Jun N-terminal Kinase (JNK) and p38 mitogen-activated protein kinase (MAPK) pathways (Figure 5d). Notably, every LD and TN sample demonstrated markedly higher expression of the gene set score (score generated from the 25 genes) without much variability between individual samples compared to the percutaneous biopsy tissues from HC, but even if compared to DM-R, early DKD, and DKD (Figure 5b).

**Figure 5.**
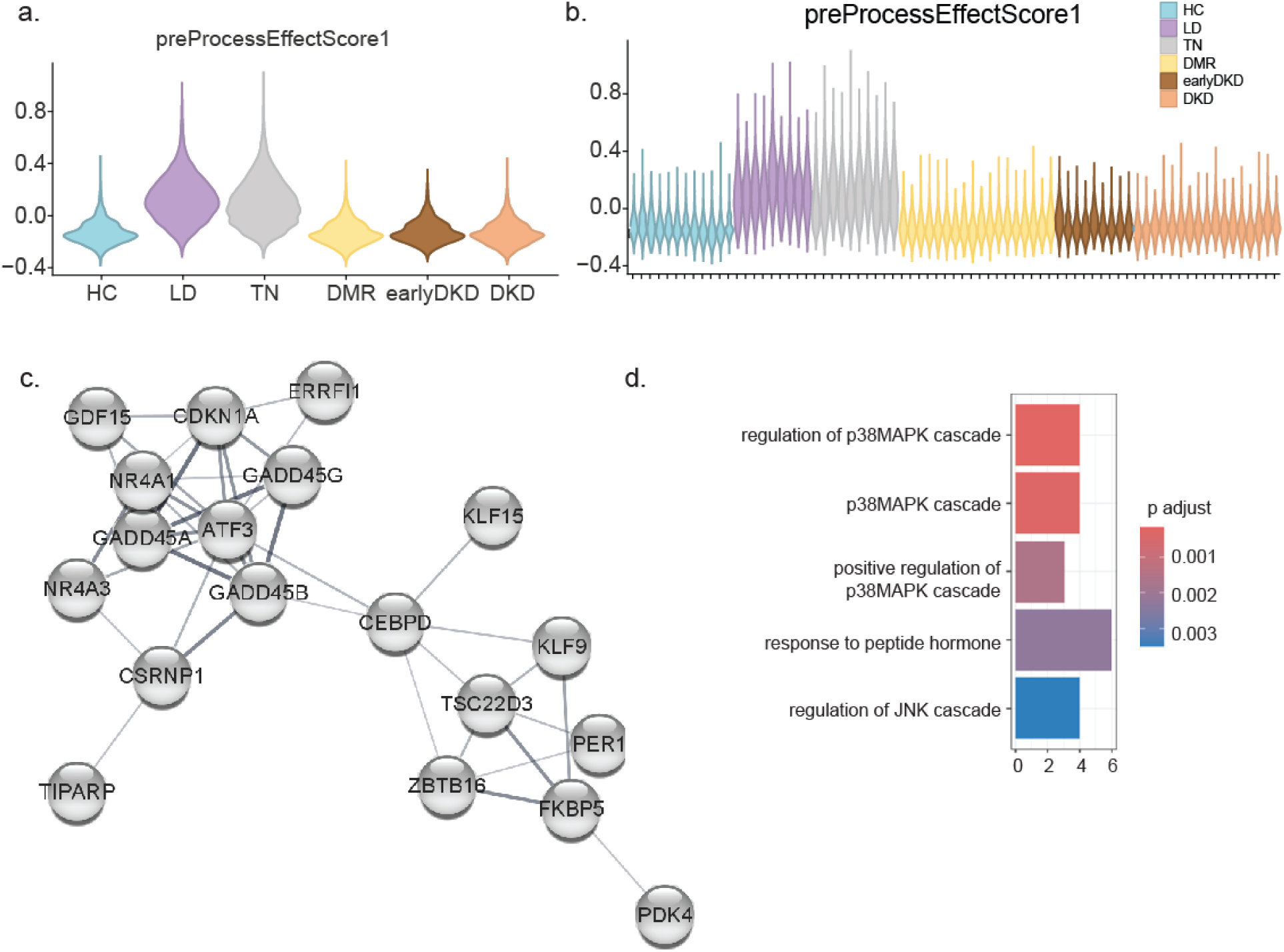
Effect of post-operative tissue procurement. Using bulk mRNA analysis, a gene set that was upregulated in post-operative tissue procurement method compared to percutaneous needle biopsies was identified. LD and TN samples were acquired by post-operative biopsy procedure and HC, DM-R, early DKD and DKD samples were by percutaneous needle biopsies. **a.** Violin density plot showing the level of the score calculated at cell level for the 25 genes that were upregulated in post-operative tissue biopsies. **d.** Violin plot showing the elevated expression of the score in every LD and TN samples compared to the needle biopsy samples. **c.** String interaction network showing direct interaction of 19/25 genes. **d.** Top 5 enriched Gene Ontology biological processes for the gene set.

### Age effect

Figure 6a shows the age distribution per sample group. HC and early DKD were considerably younger than all other sample groups. Using Pearson correlation analysis of pseudobulk read counts from both PT and aPT cells across 27 LD samples, we identified 43 genes positively associated and 41 genes negatively associated with participant age (Supplementary Table 3). Cell-level Age_Gene_Scores were calculated from single-cell mRNA expression data for these age-correlated genes (Figure 6b). Genes inversely correlated with age were more highly enriched in aPT cells. Figures 6c and 6e show positive and negative correlations between age and mean sample-level Age_Gene_Scores (positive: p<0.0002; negative: p<0.0005). Top gene ontology biological processes enriched for positively correlated genes included response to endothelium reticulum stress, negative regulation of TGFB signaling, apoptotic process, carbohydrate metabolism and response to reactive oxygen species. The genes lost with aging were enriched for positive regulation of substrate adhesion, negative regulation of serine/threonine kinase activity and intracellular signal transduction (Figure 6d, 6f).

**Figure 6.**
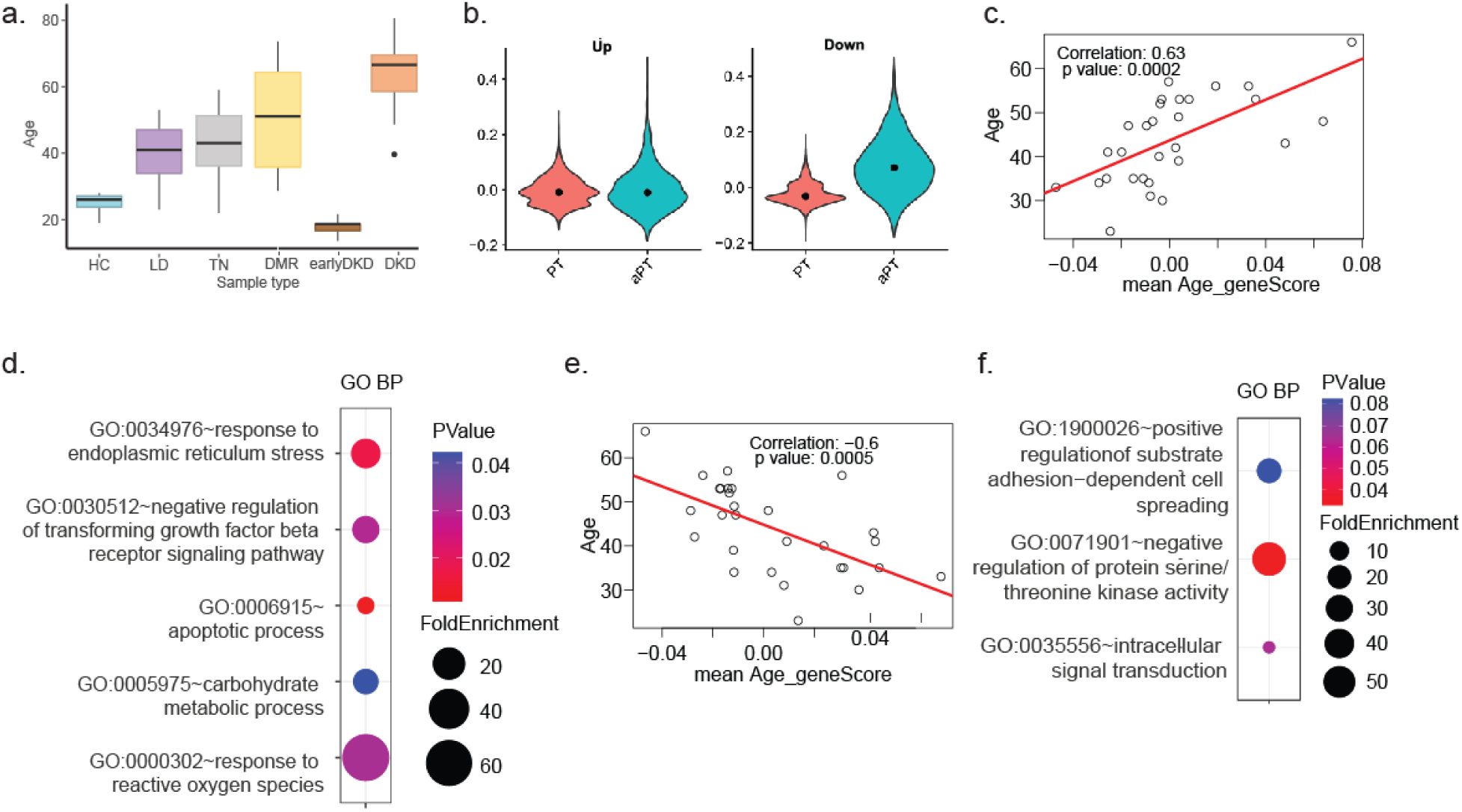
Effect of age. **a.** Bar plot showing the age distribution in each of the 6 sample types. earlyDKD and HC groups were much younger compared to other groups. **b.** Violin density plot showing the module scores calculated for the genes expressed in the proximal cells that are positively and negatively associated with age. The gene scores from the negatively associated genes were enriched in aPT cells and the gene scores from the positively associated genes did not show much differene between PT and aPT cell states. **c.** Plot showing the significant correlation between the scores from the positively age-associated genes and age of the living donors. **d.** Top enriched Gene Ontology biological processes for the positively age-associated gene set. **e.** Plot showing the significant correlation between the scores from the negatively age-associated genes and age of the living donors. **f.** Top enriched Gene Ontology biological processes for the negatively age-associated gene set.

### Pathway analysis

Hierarchical clustering of aggregated pseudo read counts at sample group level without adjusting for confounding factors is shown in Figure 7a. After correcting the effect of the confounding factors age and tissue procurement, the two diseased tissue, DKD and early DKD were grouped together, with the DM-R clustered with the reference groups.

**Figure 7.**
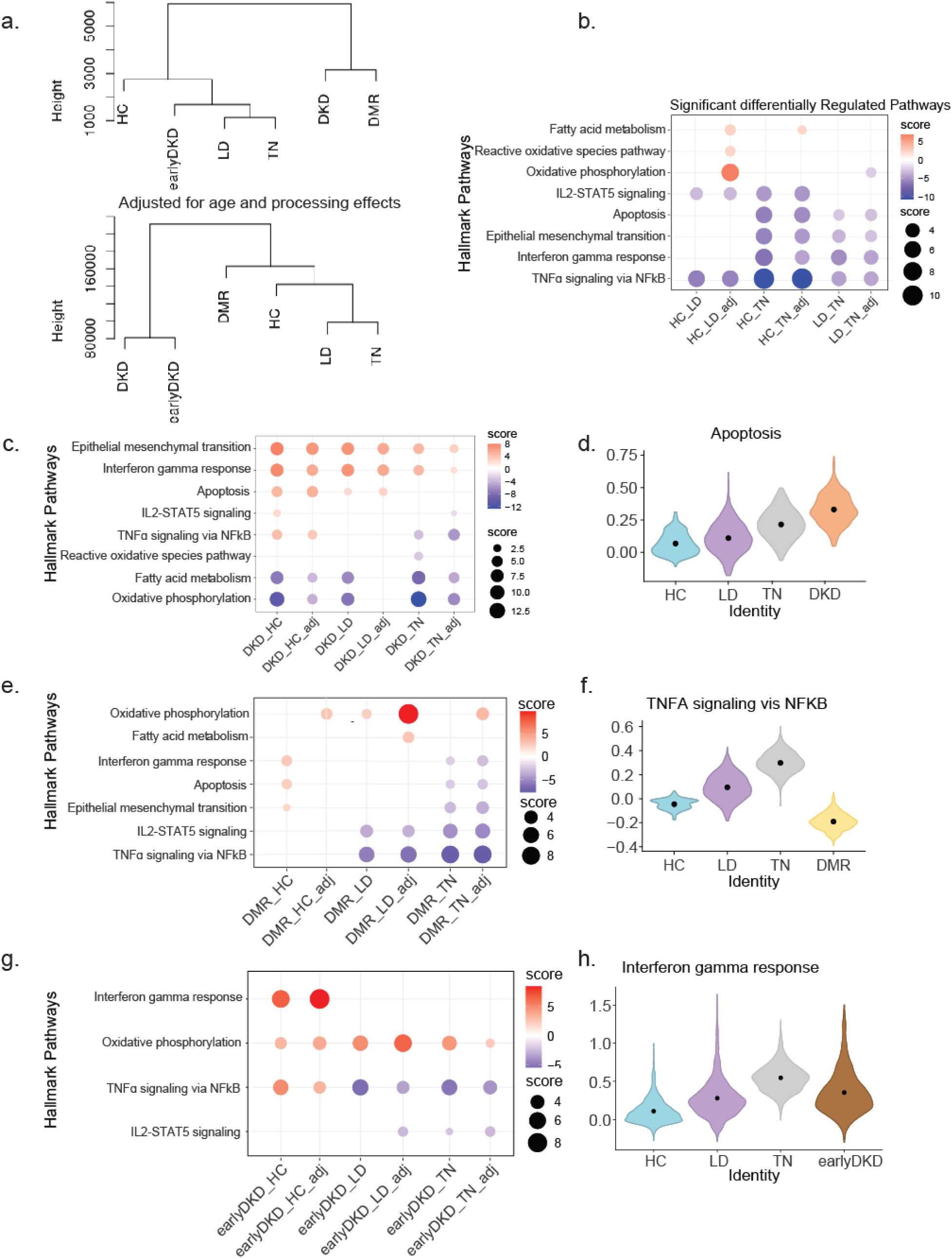
Analysis of the top pathways differentially regulated between the reference groups. **a.** Hierarchical clustering on the right was generated using aggregate read counts at the sample type level. In this analysis, DKD and DM-R clustered together, separate from the other groups. Hierarchical clustering on the left utilized aggregate read counts with batch effect correction applied via the edgeR package. For HC, earlyDKD, DM-R, and DKD, age was used as the covariate in batch effect correction; for TN and LD, both age and process effect score served as covariates. DKD and early DKD clustered together**. b** Plot displaying the top significant MSigDB Hallmark pathways (p< 0.05) associated with differentially expressed genes (p< 0.05) among the three reference groups. Differential expression analysis was performed using DESeq2, both with and without adjustment for confounding variables (age and processing effect). **c.** Plot displaying the top significant MSigDB Hallmark pathways (p< 0.05) associated with differentially expressed genes (p< 0.05) between DKD and each of the three reference groups. Differential expression analysis was performed using DESeq2, both with and without adjustment for confounding variables: age was used as the confounder for the DKD vs. HC comparison, while both age and processing effect were included for DKD vs. LD and DKD vs. TN analyses. Epithelial to mesenchymal transition is enriched in all DKD – reference groups comparisons. **d.** Violin plot showing Hallmark Apoptosis pathway scores for spots annotated as PT and aPT in the Visium transcriptomic data from DKD, HC, LD, and TN groups. The plots for DKD and TN did not show much difference. **e.** Plot displaying the top significant MSigDB Hallmark pathways (p< 0.05) associated with differentially expressed genes (p< 0.05) between DM-R and each of the three reference groups. Differential expression analysis was performed using DESeq2, both with and without adjustment for confounding variables: age was used as the confounder for the DM-R vs. HC comparison, while both age and processing effect were included for DM-R vs. LD and DM-R vs. TN analyses. DM-R showed lower enrichment for TNFA signaling viia NFKA compared to LD and TN in both with and without adjusting for confounders analyses. **f.** Violin plot showing Hallmark TNFA signaling via NFKB pathway scores for spots annotated as PT and aPT in the Visium transcriptomic data from DM-R, HC, LD, and TN groups. LD and TN spots have higher scores than HC and DM-R. **g.** Plot displaying the top significant MSigDB Hallmark pathways (p< 0.05) associated with differentially expressed genes (p< 0.05) between earlyDKD and each of the three reference groups. Differential expression analysis was performed using DESeq2, both with and without adjustment for confounding variables: age was used as the confounder for the earlyDKD vs. HC comparison, while both age and processing effect were included for earlyDKD vs. LD and early DKD vs. TN analyses. DM-R showed lower enrichment for TNFA signaling via NFKA compared to LD and TN in both with and without adjusting for confounders analyses. **h.** Violin plot displaying Hallmark Interferon Gamma response scores for PT and aPT-annotated spots in Visium transcriptomic data from early DKD, HC, LD, and TN groups. Scores were very low in HC, while LD and TN groups showed considerable variation in their distributions.

#### Differential gene expression among reference samples

To define the molecular programs characteristic of the different control tissues, we performed pathway enrichment analysis on significantly differentially expressed genes (p < 0.05) among the three reference groups. Differential expression analyses were conducted both with and without adjustments for age and procurement effects. Figure 7b presents a heatmap of pathway scores for the top Hallmark pathways from the Molecular Signatures Database (MSigDB) associated with the binary group comparisons. LD and TN compared to HC showed enrichment of TNFA signaling via NFKB and IL2-STAT5 signaling pathways, even after adjusting for confounding factors. In HC compared to LD and TN, Fatty acid metabolism was enriched following these adjustments (Figure 7b). This finding is further supported by gene expression patterns derived from Visium spatial transcriptomic profiles, where genes involved in fatty acid metabolism were highest in HC compared to LD and TN (Supplementary Figure 4). In TN compared to both LD and HC, apoptosis, epithelial-to-mesenchymal transition (EMT), and interferon gamma response showed enrichment, with same trend observed in the Visium data (Supplementary Figure 5). In aggregate the pathway findings align well with the individual stress markers profiled, with the overall highest tissue differentiation and lowest stress-inflammation profiles in HC, with substantial stress and damage pathways present even in LD biopsies and certainly in TN.

#### Differential gene expression among reference versus DKD samples

We next evaluated the impact of the choice of reference tissue on the differential gene expression output in a reference to disease comparison.

Figure 7c shows Hallmark pathway enrichment for genes significantly expressed in DKD compared to reference groups (p<0.05). Oxidative phosphorylation and fatty acid metabolism were reduced in DKD if compared to HC and TN, regardless of adjustment for confounding factors, but no such enrichment was observed in DKD versus LD after adjusting for confounders. Apoptosis was elevated in DKD compared to HC and LD but showed no significant difference from TN. Similar trend in apoptosis was observed in Visium data where Hallmark apoptosis gene scores were visualized by density plots (Figure 7d). Epithelial-to-mesenchymal transition and interferon gamma response were enriched in DKD across all reference groups (Supplementary Figure 5), while TNF signaling via NFKB was elevated in DKD if compared to HC, but decreased compared to TN, showing the impact of the reference tissue choice for this key inflammatory pathway in DKD.

Figure 7e illustrates Hallmark pathway enrichment for genes significantly expressed between DM-R and reference groups. After adjusting for confounders, oxidative phosphorylation was significantly higher in DM-R than in HC, LD, or TN, with the greatest increase observed relative to LD. Apoptosis and interferon gamma response enrichment in DM-R versus HC was no longer evident after adjusting for confounding factors, age, and procurement effects. IL2-STAT5 and TNFA signaling via NFKB remained consistently lower in DM-R compared to LD and TN, regardless of adjustments. Density plots of TNFA signaling via NFKB scores in Visium data further support higher levels in LD and TN compared to DM-R (Figure 7f).

Figure 7g displays the hallmark pathway enrichment for genes significantly expressed between early DKD and reference groups. Oxidative phosphorylation was consistently upregulated in early DKD compared to all three reference groups, regardless of confounder adjustment. The interferon gamma response was found only enriched if early DKD was compared to healthy controls (HC), but not in comparison with LD or TN. TNFA signaling was upregulated in early DKD versus HC, while the opposite trend was again observed in comparison with LD and TN. IL2-STAT5 signaling was enriched in TN compared to early DKD. Figure 7h depicts the density plot for the Interferon Gamma Response scores in Visium transcriptomic data from early DKD, HC, LD, and TN groups. Scores were lowest in HC, whereas LD and TN groups did not exhibit much variation from that of early DKD.

## DISCUSSION

Reference samples are essential in disease pathogenesis studies, yet obtaining prestine, healthy human kidney tissue is particularly challenging. Consequently, researchers have relied on kidney reference sources that do not closely match disease samples in age or procurement protocols. To evaluate the impact of these factors on differential gene expression analyses, we compared the molecular profiles of reference samples from three distinct kidney tissue sources by integrating single-cell transcriptomic data from healthy controls (HC), living donors (LD), and tumor nephrectomy (TN) samples. Although the samples originated from different projects, all study cohorts used identical tissue dissociation and single-cell RNA sequencing protocols performed by the same KPMP Tissue Interrogation Site personnel, enabling integration without batch effects.

Kidney tissue procurement, handling, and dissociation can induce early stress responses and transcriptional changes(12). However, since all samples in this study were stored and processed using an identical protocol, observed differences are likely driven by procurement methods or donors’ physiological and clinical characteristics. Samples were obtained via percutaneous needle research biopsy (HC, early DKD, DM-R, DKD) or post-operative biopsy (LD, TN). HC samples displayed lower mitochondrial read content per cell and reduced expression of early stress markers compared to LD and TN, suggesting superior tissue viability with the percutaneous needle biopsy procurement (Figure 3a). Additionally, a gene set associated with post-operative procurement was consistently elevated in all LD and TN samples, with enriched pathways involving stress-related JNK and p38MAPK cascades that exceeded those seen in DKD biopsy tissues (Figure 5).

Healthy control samples had minimal or absent expression of disease markers such as IL18, LCN2, HAVCR1, and VCAM1, underscoring their pristine quality (Figure 3c & 3d). Lower levels of kidney damage markers and B cell infiltration in HC may be attributed to both the biopsy procedure and the younger age of these donors compared to LD and TN.

LD samples exhibited higher expression of HAVCR1, which encodes Kidney Injury Marker 1 (KIM1), compared to HC and TN (adjusted p< 0.05). HAVCR1 was significantly increased in the adaptive/maladaptive proximal cell state, which was prevalent in LD and TN (Supplementary Figure 3). Its expression was increased in DKD compared to LD, was unchanged in DM-R versus LD, and was significantly lower in early DKD versus LD, suggesting an age-related increase confounding the DKD effect on in this comparison, consistent with age-related increases in KIM1 observed in blood DKD biomarker studies (13).

TN samples consisted of non-cancerous tissue from donors with high eGFR (>80), minimal pathology, and no diabetes or hypertension. IL18, an early CKD marker and pro-inflammatory cytokine(14), was significantly elevated in TN compared to LD and HC. High enrichment of apoptosis, EMT, and Interferon gamma response in TN compared to other reference groups (Supplementary Figure 5) observed in the Visium data further support the underlying stress signal present in TN samples, despite the fact that only clinical and structural normal kidney tissue was used for these analyses.

We identified an age-associated gene set expressed in proximal cells, with age-correlated genes enriched for endoplasmic reticulum stress, apoptosis, and reactive oxygen species response processes linked to aging(15–17). Conversely, genes negatively correlated with age were enriched for substrate adhesion, negative regulation of serine/threonine kinase activity, and intracellular signal transduction, all known to decline with age(18–20).

As confounding factors can substantially impact transcriptomic results(21), we also reported pathway enrichment after adjusting for age and procurement effects on gene expression profiles. This approach allowed to correct for some of these effects intrinsic to easily obtainable TN and LD reference tissue, as seen for fatty acid metabolism, reactive oxygen species, and oxidative phosphorylation in HC compared to LD (Figure 7b).

The DM-R group, despite longstanding disease and risk exposure, showed minimal renal impairment, revealing potential protective mechanisms against diabetic kidney injury. While the enrichment of interferon gamma response and apoptosis in DM-R versus HC diminished after accounting for confounders (Figure 7d), oxidative phosphorylation remained consistently elevated in DM-R compared to reference groups, most likely reflective of the higher metabolic demand in the diabetic state, as this was also observed in the early DKD kidneys(22, 23) (24).

Selection of reference tissue also impacted the detection of inflammatory pathways in DKD. TNF-alpha is well known to activate NF-κB via TNF receptor 1 (TNFR1), leading to transcription of target genes implicated in DKD(25). Notably, increased TNF signaling via NF-κB was only visible in early DKD when compared with HC. If early DKD was compared to LD or TN a significant reduction in TNF signaling in early DKD was found, even when adjusting for confounders, highlighting the critical role of the comparator selection for differential gene expression analyses.

However, some pathways remained consistently higher in all DKD versus reference group comparisons, even after adjusting for confounders, including EMT and interferon gamma response known to be robustly elevated in diabetic kidney disease (26, 27).

A limitation of this study is the small sample size per group in each category, precluding adjustments for additional confounders in lower abundance cell types and, most notably to match gender distributions(28).

In summary, our findings demonstrate the critical role of references tissue sources and procurement methods for comparative gene expression analysis. In particular, the inclusion of healthy volunteer research biopsies from young adults allowed us to anchor our analyses to optimal kidney tissue. Although recruiting healthy control (HC) volunteers is challenging, percutaneous needle biopsies from HCs are molecularly pristine and ideal for studying biological processes across DKD stages, particularly when early or discreate disease changes are being mapped. However, when comparing disease biopsies from older patients, age must be considered as a confounding factor, and aged match controls should be considered, based on experimental questions to be addressed (i.e., exposure to differential environmental factors over the lifespan). Living donor (LD) and tumor-nephrectomy (TN) samples obtained through post-surgical biopsies are more accessible and have resulted in larger cohorts available to be studied. However, substantial procurement stress responses often impact the same pathways also altered in the disease processes of interest, able to obscure the disease signal. As shown in this study, confounding effects can, to some extent, be corrected by adjustment for age and procurement factors. At a minimum, the data presented in this study will allow researchers to flag potential competing activation in both disease and reference samples in an analysis set. Finally, our study justifies the development of a framework for sufficiently diverse healthy tissue reference datasets for future analyses (29).

## MATERIALS & METHODS

### Sex as a biological variable

Both male and female participants who were enrolled in all the cohorts were included in the analyses.

### Human data

Key clinical and demographic characteristics of participants who contributed kidney biopsy samples in the three reference and three diabetes-related groups are summarized in Table 1. For this study, we have used data from kidney tissue collected by the Kidney Precision Medicine Project (KPMP, details see KPMP.org), which focuses on molecular mechanisms in acute kidney injury (AKI) and chronic kidney disease (CKD). From the KPMP cohort, nine LD, seventeen DKD, and eighteen DM-R biopsy samples were used in this study. LD and TN samples were obtained through post-operative surgical biopsies, with LD biopsies performed on the explanted kidney before implantation into the transplant recipient. HC and diabetic samples were collected via percutaneous research needle biopsies.

**Table 1:** Characteristics of study participants.

| Sample category | Samples (N) | Average Age in years ± standard deviation | Sex % Female (Male) | Race (%) | | | |
| --- | --- | --- | --- | --- | --- | --- | --- |
|  |  |  |  | Asian | Black | White | Other<sup>a</sup> |
| Healthy volunteer (HC) | 12 | 25.5 ± 3 | 46 (54) | 8 | 0 | 91 | 0 |
| Preperfusion living donor biopsy (LD) | 9 | 37.8 ± 10.4 | 77 (23) | 11 | 0 | 78 | 11 |
| Tumor-nephrectomy (TN) | 9 | 42.7 ± 11.67 | 33 (67) | 0 | 0 | 100 | 0 |
| Diabetic mellitus-resilient (DM-R) | 18 | 50.05 ± 14.52 | 61 (39) | 0 | 0 | 94 | 6 |
| Early chronic kidney disease (early DKD) | 9 | 17.09 ± 2.29 | 44 (56) | 0 | 22 | 67 | 11 |
| Diabetic kidney disease (DKD) | 17 | 60.46 ± 10.53 | 65 (35) | 6 | 35 | 47 | 12 |

<sup>a</sup>Native American/Alaskan, Pacific Islanders/Hawaiian, not known

The TN samples comprised tumor-free kidney cortical tissue from nephrectomy specimens rapidly sourced in the operating room for molecular analyses (PRECISE cohort, University of Michigan (30). The 9 samples were selected from donors with high eGFR (>80), low pathology, no diabetes and no hypertension.

The 9 early DKD samples were from the Impact of Metabolic Surgery on Pancreatic, Renal and Cardiovascular Health in Youth with Type 2 Diabetes (IMPROVE-T2D) study and the Renal Hemodynamics, Energetics and Insulin Resistance in Youth Onset Type 2 Diabetes Study (Renal HEIR) and 12 healthy control samples (HC) from research kidney biopsies performed on healthy volunteers in the Control of Renal Oxygen Consumption, Mitochondrial Dysfunction, and Insulin Resistance study (CROCODILE), as previously described (23).

For identifying the genes associated with age using weighted correlation network analysis, (WGCNA), we used data from tissue obtained from living kidney transplant donors (LD). In addition to the 9 KPMP LD samples, we included 18 samples from the University of Michigan living donor cohort generated for the Human Kidney Transplant Transcriptomic Atlas.

After procurement with identical single cell RNA-seq protocols all samples were processed by the KPMP Premier tissue interrogation core facility staff of University of Michigan.

### Sex as a biological variable

All cohorts used in the study include sex as a key variable in recruitment to ensure representative study population.

### Single cell data generation

Single cell dissociation of the samples was performed using Liberase TL at 37°C. Detailed sample processing protocol can be found in protocol.io: dx.doi.org/10.17504/protocols.io.7dthi6n. The single cell suspension was immediately transferred to the University of Michigan Advanced Genomics Core facility for further processing. 10X-Genomics single cell RNA sequencing (scRNAseq) of the samples was done according to dx.doi.org/10.17504/protocols.io. The raw sequences files were aligned to the reference genome (GRCh38 (hg38)) and the feature-barcode matrices were generated by Cell Ranger program embedded in 10X-Genomics analysis software.

### Single cell data analysis

The read count matrices were initially processed using SoupX (v1.5.0) to correct for ambient mRNA contamination. Data processing was performed using the Seurat R package (version 5.0). Quality control was enforced with cutoffs of >500 and <5000 genes per cell, and <50% mitochondrial reads per cell. Each sample group underwent normalization, variable gene identification, and principal component analysis independently. Subsequently, the integration of all six sample types was achieved via reciprocal principal component analysis (RPCA).

Dimensionality reduction using Uniform Manifold Approximation and Projection (UMAP) and unsupervised clustering (resolution 0.4) were conducted on the integrated dataset. Cell clusters were annotated based on markers of major nephron segments, as well as endothelial, interstitial, and immune cell types. Further analyses were performed on a down-sampled integrated object containing 20,000 randomly selected cells from each sample type (the integrated dataset containing a total of 120,000 cells from all 6 sample types together).

### Post-operative tissue procurement effect analysis

The gene set upregulated in post-operative sample types (LD and TN) compared to other groups (HC, DM-R, early DKD, and DKD) was identified via differential expression analysis using edgeR (31) on pseudobulk RNA read counts. The final gene set was selected after manually confirming overexpression across all samples in LD and TN groups to avoid sample-to-sample variation. The interaction network of this gene set was constructed using STRING (11) and visualized as a network in Cytoscape(32). Additionally, gene ontology biological processes enrichment analysis for this gene set was conducted using the ClusterProfiler R package(33).

### Age effect

To evaluate the impact of age within a reference group while minimizing confounding factors, we performed a correlation analysis between gene expression and donor age using 27 living donor samples, as the HC and TN groups were underpowered. We identified the top 2,500 highly variable genes in proximal cells from normalized pseudo read counts (aggregated by sample and processed with edgeR (31) and assessed their correlation with age using Pearson’s correlation test. For the positively and negatively age-correlated genes, we calculated an “Age_Gene_Score” in the integrated Seurat object using the AddModuleScore function.

### Adjusting for procurement and age effect in differential expression analyses

Differential expression analyses were performed between disease and reference groups using the DESeq2 (34) R package on pseudo-bulk mRNA counts, generated by aggregating raw read counts from proximal cells for each sample. Analyses were conducted both unadjusted and adjusted for age and procurement effect as confounding factors. Each of the three disease groups was separately compared with each of the three reference groups. To account for procurement effect, we calculated a composite score by averaging Z-scores of 25 genes positively associated with post-operative tissue procurement. Participant age at biopsy was included as a covariate in the adjusted models.

### Pathway enrichment analysis

Pathway activity enrichment was assessed using decoupleR(35) on genes with significant differential expression (p < 0.05), as identified by DESeq2 from aggregated pseudo read counts comparing proximal cells in reference and disease groups. Hallmark pathways from the Molecular Signatures Database (MSigDB) were used as the resource for enrichment analysis. Pathways with enrichment p-values below 0.05 were deemed significant.

### Validation of pathway activity using Visium transcriptomic data

To assess pathway enrichment of differentially expressed genes between disease and reference groups, we analyzed transcriptomic datasets generated using 10X-Genomics Visium technology. Data for LD, TN, DM-R, and DKD were obtained from the KPMP repository (https://atlas.kpmp.org/repository/), while HC and early DKD datasets were generated locally for an unrelated project. Human kidney tissue was prepared and imaged according to the Visium Spatial Gene Expression (10x Genomics) manufacturer’s protocol (CG000240, Visium Tissue Preparation Guide) and as previously described (1, 36).

Each group included three samples, except tumor-nephrectomy (TN), which had two. Due to the limited sample size per group, all Visium based validations were conducted on the pathway enrichment analyses from single-cell data without confounder adjustment.

Data integration was performed using Seurat. Briefly, each dataset underwent Single-Cell Transform (SCT) normalization, followed by dimensionality reduction with principal component analysis (PCA). Integration anchors were identified using reciprocal PCA (RPCA), and the datasets were merged into a single integrated object. Additional dimensionality reduction was then performed on the integrated object using both PCA and UMAP. Spot annotation was carried out with Seurat’s label transfer method, using the integrated and down-sampled single-cell dataset as the reference. For the validation of the observations from the proximal single cell data, we focused on the Visium spots annotated as proximal (PT and aPT).

### Statistics

To identify preprocessing effect genes, we first used Seurat’s FindMarkers function to detect genes that were differentially expressed in LD and TN compared to other groups, applying an adjusted p-value threshold of < 0.05. Next, edgeR was performed on the pseudobulk read counts of these significant genes. Genes with a p-value < 0.05 and a logFC > 0 were then manually validated and classified as preprocessing effect genes.

Pathway activity enrichment was assessed using decoupleR on genes with significant differential expression (p < 0.05), as identified by DESeq2 from aggregated pseudo read counts comparing proximal cells in reference and disease groups. Pathways with enrichment p-values below 0.05 were considered as significant.

### Study approval

All samples used in this study were obtained with patient consent and with the approval of IRBs of participating institutions, described below. The KPMP study is reviewed and approved under a protocol by the KPMP single IRB of the University of Washington Institutional Review Board (IRB 20190213). The PRECISE cohort and the Human Kidney Transplant Transcriptomic Atlas studies were approved by the IRB of the University of Michigan. CROCODILE, Renal HEIR, and IMPROVE-T2D, were approved by the Colorado Multiple Institutional Review Board (COMIRB). Written, informed consent was received from each participant and/or parents as appropriate for age prior to enrollment.

## Data availability

The KPMP single cell data used in this study is available from https://atlas.kpmp.org/repository/. The single cell count matrices for the Human Kidney Transplant Transcriptomic Atlas is available from the Gene Expression Omnibus under accession number GSE 169285. Healthy control and early chronic disease sample read count matrices are available at GSE220939, and GSE279086. The tumor-nephrectomy data can be found in https://cellxgene.cziscience.com/collections/a98b828a-622a-483a-80e0-15703678befd.

## Author Contributions

RM designed the study, analyzed data, interpreted results, and drafted the manuscript. MK designed the study, interpreted results, and reviewed and edited the manuscript. EO and BG performed the single cell experiments. PB, CC, MB, JBH, RMF and MTE provided transcriptomic data, and interpreted the results. YC, LP, PL, PB, MS, ESW, JS, SER, SW and AN provided the kidney tissue samples used in the study. PK, LS, EO, CC, BG, CS, FA, and CB edited the manuscript. All authors reviewed the manuscript, contributed to revising the manuscript and approved the final version.

## Supporting information

Supplemental Figures and acknowledgement

## Acknowledgement

We are deeply indebted to the generosity of participants volunteering to donate tissue primarily for research purposes despite receiving no direct immediate benefit to their clinical care. We thank the clinical coordinators for their efforts in participant enrolments and biopsy tissue procurement.

