## Supplemental Figures and acknowledgement for "Not all controls are made equal: Definition of human kidney reference samples by single cell gene expression profiles"

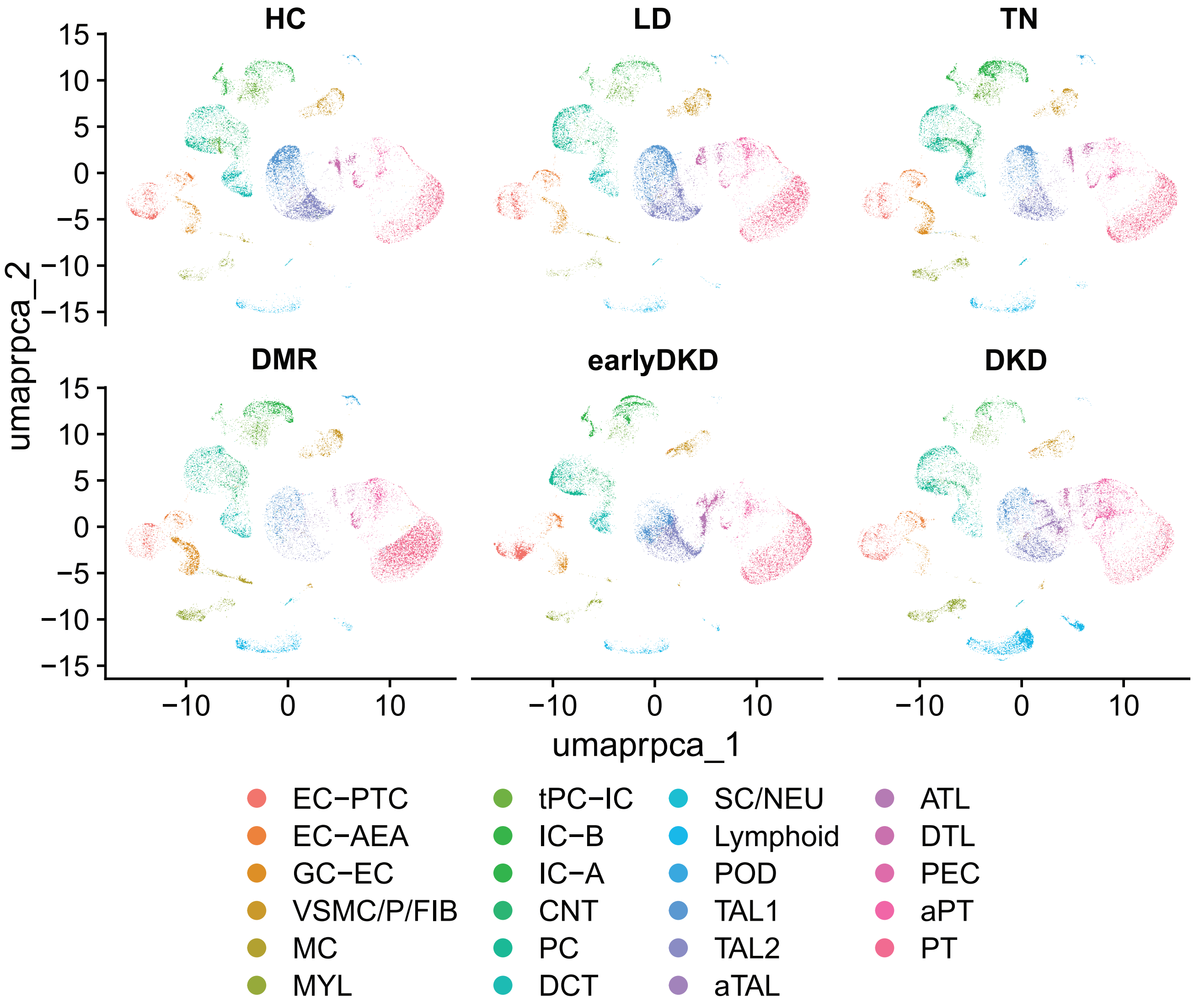


**Supplementary Figure S1. Split UMAP of the 6 sample groups studied**
Abbreviations: HC- healthy controls; LD- living kidney donor; TN- tumor nephrectomy; DMR - diabetes mellitus resistor; DKD - diabetic kidney disease;
Cell types: podocyte, POD; parietal epithelial cell, PEC; proximal tubule, PT; adaptive/maladaptive, aPT; descending thin loop of Henle, DTL; ascending thin loop of Henle, ATL; thick ascending loop of Henle, TAL; adaptive/maladaptive thick ascending loop of Henle, aTAL; distal convoluted tubule, DCT; connecting tubule, CNT; principal cell, PC; intercalated type A, IC-A; Intercalated type B, IC-B; transient between PC and IC, tPC-IC; endothelial cell EC; efferent and afferent arteriolar endothelial cells EC-AEA; glomerular endothelial cell, EC-GC; fibroblast, FIB; vascular smooth muscle cell, VSMC; pericyte, P; mesangial cell, MC; myeloid cell, MYL

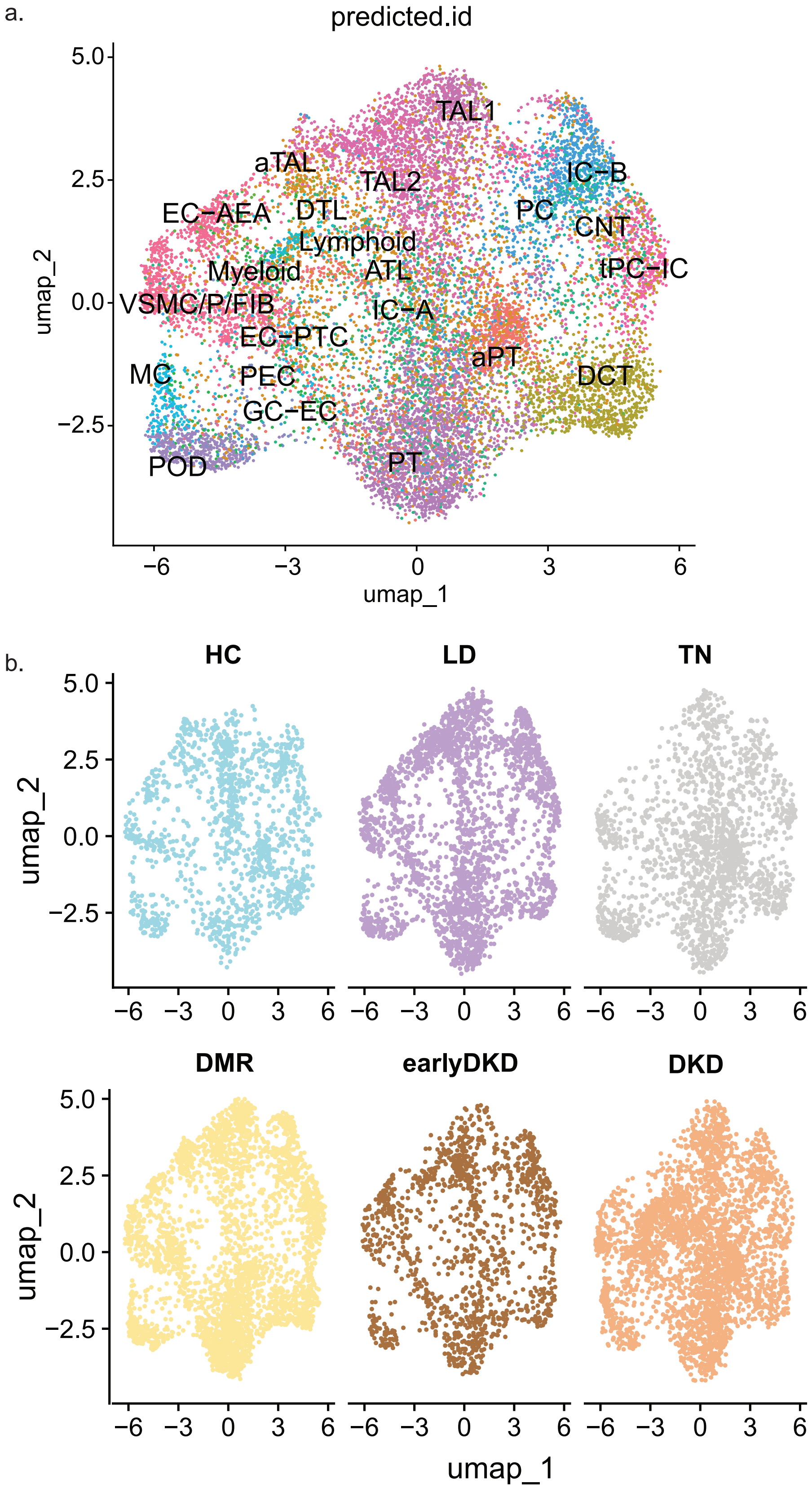


**Supplementary Figure S2. Visium data UMAPs.** Combined (a) and split (b) UMAPs of the integrated Visium dataset. 17 samples were used in the integration analysis, three samples per HC, LD, DM-R, early DKD, and DKD; two samples in TN. Abbreviations: HC- healthy controls; LD- living kidney donor; TN- tumor nephrectomy;DMR - diabetes mellitus resistor; DKD - diabetic kidney disease.

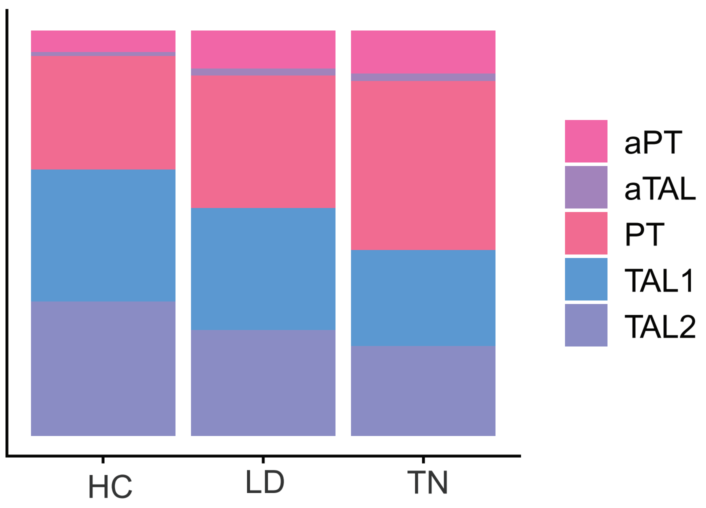


**Supplementary Figure S3: Proportion of proximal tubule and thick ascending loop cell states in the reference groups.**

Abbreviations: HC- healthy controls; LD- living kidney donor; TN- tumor nephrectomy; PT-proximal tubule, adaptive/maladaptive proximal tubule - aPT; TAL-thick ascending loop of Henle; adaptive/maladaptive thick ascending loop of Henle-aTAL.

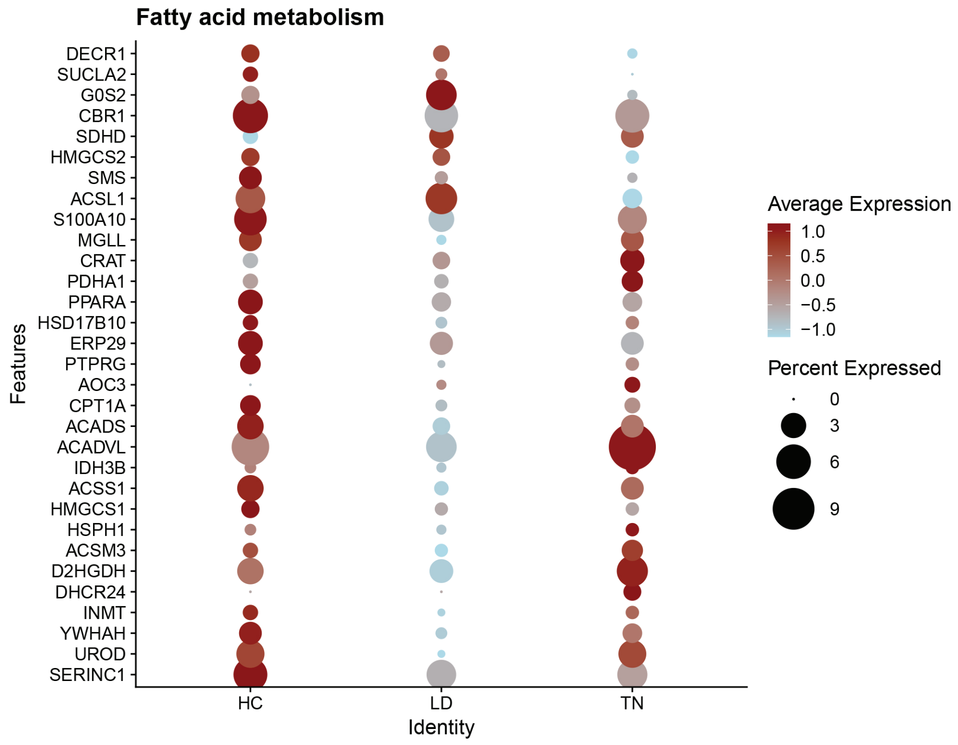


**Supplementary Figure S4: Expression of genes enriched for fatty acid metabolism in the proximal cells of Visium data from the three reference groups.**

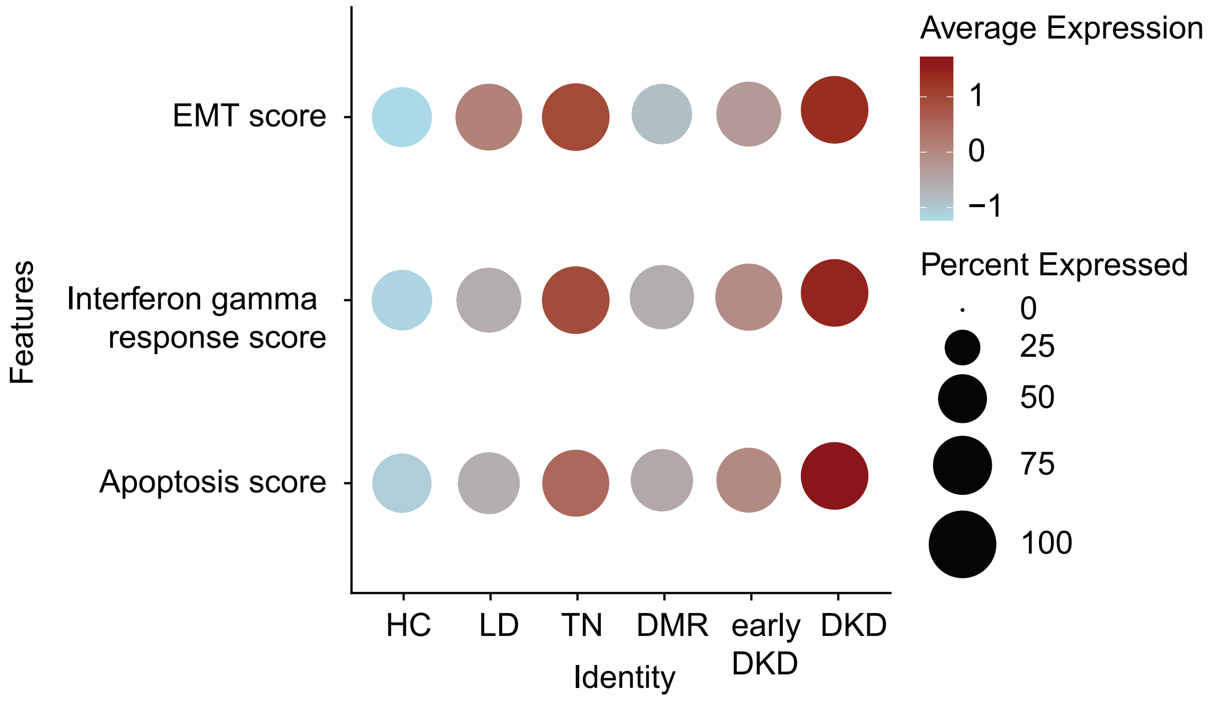


**Supplementary Figure S5. Pathway scores in proximal tubule cells across all groups.** Dot plot showing the scores generated from the genes enriched for apoptosis, interferon gamma response and epithelial to mesenchymal response (EMT) in the Visium data of proximal cells from all sample groups studied.

### Supplementary Acknowledgements


Kidney Precision Medicine Project (KPMP) members

| Name | Affiliated Institution | Role |
|---|---|---|
| Alexander Morales | Beth Israel Deaconess Medical Center | Biopsy Operator |
| Mark E. Williams | Beth Israel Deaconess Medical Center | Co-Investigator |
| Stewart H. Lecker | Beth Israel Deaconess Medical Center | Co-Investigator,Biopsy Operator |
| Laurence H Beck Jr | Boston Medical Center | Co-Investigator |
| IM Schmidt | Boston Medical Center | Co-Investigator |
| Ashish Upadhyay | Boston Medical Center | Co-Investigator |
| Ashish Verma | Boston Medical Center | Co-Investigator |
| Minxin Lu | Boston Medical Center | Co-Investigator,Study staff |
| Yan Zhou | Boston Medical Center | Fellow |
| Stephanie J. Aw | Boston Medical Center | Other |
| Kifle Gebre | Boston Medical Center | Other |
| Shana Maikhor | Boston Medical Center | Other |
| Keyvona Moultrie | Boston Medical Center | Other |
| Florencia A. Rojas-Miguez | Boston Medical Center | Other |
| Molly C Geraghty | Boston Medical Center | Pathologist |
| Joel M Henderson | Boston Medical Center | Pathologist |
| Narasimhan, R. | Boston Medical Center | Pathologist |
| Sushrut S. Waikar | Boston Medical Center | Principal Investigator |
| Marie Florence Calixte | Boston Medical Center | Research Coordinator |
| Courtney Huynh | Boston Medical Center | Research Coordinator |
| Astrid Larson | Boston Medical Center | Research Coordinator |
| Ingrid F Onul | Boston Medical Center | Research Coordinator |
| Pranav Yadati | Boston Medical Center | Research Coordinator |
| Guanghao Yu, BA | Boston Medical Center | Research Coordinator |
| M. Todd Valerius | Brigham and Women's Hospital | Co-Investigator |
| Gearoid Michael McMahon | Brigham and Women's Hospital | Co-Investigator,Biopsy Operator |
| Mia R. Colona | Brigham and Women's Hospital | Other |
| Helmut Rennke | Brigham and Women's Hospital | Other |
| Astrid Weins | Brigham and Women's Hospital | Pathologist |
| Anna Greka | Broad Institute | Other |
| Jamie L. Marshall | Broad Institute | Other |
| Nir Hacohen | Broad Institute | Principal Investigator |
| Mark P. Aulisio | Case Western Reserve University | Co-Investigator |
| William S. Bush | Case Western Reserve University | Co-Investigator |
| Dana C. Crawford | Case Western Reserve University | Co-Investigator |
| Crystal A Gadegbeku | Cleveland Clinic | Co-Investigator |

| | | |
|---|---|---|
| Jonathan J Taliercio | Cleveland Clinic | Co-Investigator |
| Charles O'Malley | Cleveland Clinic | Co-Investigator,Biopsy Operator |
| Leslie Cooperman | Cleveland Clinic | Other |
| Stacey Jolly | Cleveland Clinic | Other |
| Ellen Palmer | Cleveland Clinic | Other |
| Leal Herlitz | Cleveland Clinic | Pathologist |
| Michael Kuperman | Cleveland Clinic | Pathologist |
| Kassandra Spates-Harden | Cleveland Clinic | Patient Partner |
| John F. O'Toole | Cleveland Clinic | Principal Investigator |
| John R. Sedor | Cleveland Clinic | Principal Investigator |
| Emilio D Poggio | Cleveland Clinic | Principal Investigator,Co-Investigator |
| Lakeshia Bush | Cleveland Clinic | Research Coordinator |
| Vivian Jeffers | Cleveland Clinic | Research Coordinator |
| Kiasha Jones | Cleveland Clinic | Research Coordinator |
| Marina Markovic | Cleveland Clinic | Research Coordinator |
| Teresa Randle | Cleveland Clinic | Research Coordinator |
| Dianna Sendrey | Cleveland Clinic | Research Coordinator |
| Paul S. Appelbaum | Columbia University | Other |
| Olivia Balderes | Columbia University | Other |
| Pietro A. Canetta | Columbia University | Other |
| Karla Mehl | Columbia University | Other |
| Vivette . D'Agati | Columbia University | Pathologist |
| Jonathan Barasch | Columbia University | Principal Investigator |
| Andrew S. Bomback | Columbia University | Principal Investigator |
| Krzysztof Kiryluk | Columbia University | Principal Investigator |
| German varela | Columbia University | Research Coordinator |
| Laura Barisoni | Duke University | Co-Investigator,Pathologist |
| Akhil Ambekar | Duke University | Fellow |
| Xiang Li | Duke University | Fellow |
| Bangchen Wang | Duke University | Pathologist |
| Theodore Alexandrov | European Molecular Biology Laboratory | Co-Investigator |
| Jia-Yun Chen | Harvard University | Co-Investigator |
| Nils Gehlenborg | Harvard University | Co-Investigator |
| Jia-Ren Lin | Harvard University | Fellow |
| Yi Zhang | Harvard University | Fellow |
| Mark S. Keller | Harvard University | Other |
| Seymour Rosen | Harvard University | Pathologist |
| Sandro Santagata | Harvard University | Principal Investigator |
| Sharon B Bledsoe | Indiana University | Biopsy Operator |
| Ying-Hua Cheng | Indiana University | Co-Investigator |

| | | |
|---|---|---|
| Kenneth W. Dunn | Indiana University | Co-Investigator |
| Michael Ferkowicz | Indiana University | Co-Investigator |
| K. J. Kelly | Indiana University | Co-Investigator |
| Ricardo Melo Ferreira | Indiana University | Co-Investigator |
| Marcelino Rivera | Indiana University | Co-Investigator |
| Timothy A. Sutton | Indiana University | Co-Investigator |
| Debora Gisch | Indiana University | Co-Investigator,Fellow |
| Azuma Nanamatsu | Indiana University | Fellow |
| Mohammad A. Sohail | Indiana University | Fellow |
| Mahla Asghari | Indiana University | Other |
| Andreas Bueckle | Indiana University | Other |
| Curtis Warfield | Indiana University | Patient Partner |
| Tarek M. El-Achkar | Indiana University | Principal Investigator |
| Pierre C. Dagher | Indiana University | Principal Investigator |
| Michael T Eadon | Indiana University | Principal Investigator |
| Danielle Janosevic | Indiana University | Principal Investigator |
| James C. Williams, Jr. | Indiana University | Principal Investigator |
| Abraham Verdoes | Indiana University | Project Manager |
| Devin M. Wright | Indiana University | Project Manager |
| Stephanie Wofford | Indiana University | Research Coordinator |
| Daria Barwinska | Indiana University | Study staff |
| William S. Bowen | Indiana University | Study staff |
| Angela R. Sabo | Indiana University | Study staff |
| Jennifer Stashevsky | Indiana University | Study staff |
| Katy Börner | Indiana University Bloomington | Other |
| Ellen M. Quardokus | Indiana University Bloomington | Other |
| Jini Ashok Bhanushali | Indiana University Bloomington | Project Manager |
| Bruce W. Herr II | Indiana University Bloomington | Project Manager,Other |
| Elizabeth G. Record | Indiana University Bloomington | Research Coordinator |
| Derek M. Fine | Johns Hopkins University | Biopsy Operator |
| C. John Sperati | Johns Hopkins University | Biopsy Operator |
| Steven Menez | Johns Hopkins University | Co-Investigator |
| Mohamed G. Atta | Johns Hopkins University | Co-Investigator,Biopsy Operator |
| Jose M Monroy-Trujillo | Johns Hopkins University | Co-Investigator,Biopsy Operator |
| Yumeng Wen | Johns Hopkins University | Co-Investigator,Fellow |
| Lauren Bernard | Johns Hopkins University | Other |
| Noralinda B. Viloria | Johns Hopkins University | Other |
| Alan Xu | Johns Hopkins University | Other |
| Avi Z. Rosenberg | Johns Hopkins University | Pathologist |
| Chirag R. Parikh | Johns Hopkins University | Principal Investigator |
| Celia P. Corona-Villalobos | Johns Hopkins University | Project Manager |
| Mitali Barik | Johns Hopkins University | Research Coordinator |

| | | |
|---|---|---|
| Maria Chilo Bejarano | Johns Hopkins University | Research Coordinator |
| Jeanine Hernandez | Johns Hopkins University | Research Coordinator |
| Sonya Shah | Johns Hopkins University | Research Coordinator |
| Ashley R. Wang | Johns Hopkins University | Research Coordinator |
| Paolo S. Silva | Joslin Diabetes Center | Co-Investigator |
| Jennifer K. Sun | Joslin Diabetes Center | Co-Investigator |
| Isabel Donohoe | Joslin Diabetes Center | Other |
| Camille Johansen | Joslin Diabetes Center | Other |
| N/A | Joslin Diabetes Center | Other |
| Rosas, Sylvia E. | Joslin Diabetes Center | Principal Investigator |
| Sophia A. Angus | Joslin Diabetes Center | Research Coordinator |
| Sarah W Chen | Joslin Diabetes Center | Research Coordinator |
| Asari Henshaw | Joslin Diabetes Center | Research Coordinator |
| Neil Roy | Joslin Diabetes Center | Research Coordinator |
| Imane H. Samari | Joslin Diabetes Center | Research Coordinator |
| Gabriel Zeinoun | Joslin Diabetes Center | Research Coordinator |
| Mallory Mandel | Joslin Diabetes Center | Study staff |
| Jenny Molina-Guzman | Joslin Diabetes Center | Study staff |
| Anna Kate Stawicki | Joslin Diabetes Center | Study staff |
| Melissa D. Rubinsky | Joslin Diabetes Center | Study staff,Other |
| Julia A. Welch | Joslin Diabetes Center | Study staff,Other |
| Lili Chan | Mount Sinai | Biopsy Operator |
| Daniel Stalbow | Mount Sinai | Biopsy Operator |
| Evren U Azeloglu | Mount Sinai | Co-Investigator |
| Jens Hansen | Mount Sinai | Co-Investigator |
| John Cijiang He | Mount Sinai | Co-Investigator |
| Carol R. Horowitz | Mount Sinai | Co-Investigator |
| Ravi Iyengar | Mount Sinai | Co-Investigator |
| Joji Tokita | Mount Sinai | Co-Investigator,Biopsy Operator |
| Patricia Kovatch | Mount Sinai | Co-Investigator,Other |
| Marina de Cos | Mount Sinai | Fellow |
| Jonathan Haydak | Mount Sinai | Fellow,Patient Partner |
| Lili Gai | Mount Sinai | Other |
| Patricia Kovatch | Mount Sinai | Other |
| Timothy D. Quinn | Mount Sinai | Other |
| Ronald E. Gordon | Mount Sinai | Pathologist |
| Ritu Gupta | Mount Sinai | Pathologist |
| Isaac E Stillman | Mount Sinai | Pathologist |
| Stephen C Ward | Mount Sinai | Pathologist |
| Kirk N Campbell | Mount Sinai | Principal Investigator |
| Steven G. Coca | Mount Sinai | Principal Investigator |
| Jonathan Himmelfarb | Mount Sinai | Principal Investigator |
| Kristin Meliambro | Mount Sinai | Principal Investigator |

| | | |
|---|---|---|
| Girish N Nadkarni | Mount Sinai | Principal Investigator |
| Mark L Green | Mount Sinai | Project Manager |
| Brandon G Larson | Mount Sinai | Project Manager |
| Lorraine Evo-Ortega | Mount Sinai | Research Coordinator |
| Gina Koch | Mount Sinai | Research Coordinator |
| Marissa Patel | Mount Sinai | Research Coordinator |
| Sean Lefferts | Mount Sinai | Research Coordinator,Project Manager |
| Tejas Rao | Mount Sinai | Study staff |
| Samuel Mon-Wei Yu | Mount Sinai | Study staff |
| Pottumarthi V Prasad | Northwestern University | Co-Investigator |
| Samir V Parikh | Ohio State University | Co-Investigator |
| Brad H. Rovin | Ohio State University | Principal Investigator |
| Gek Cher Chan | Other | Co-Investigator |
| Yijiang Chen | Other | Co-Investigator |
| Joana P. Gonçalves | Other | Co-Investigator |
| Andrew Janowczyk | Other | Co-Investigator |
| Blue B. Lake | Other | Co-Investigator |
| Roy Lardenoije | Other | Co-Investigator |
| Rosamond Rhodes | Other | Co-Investigator |
| Milda R. Saunders | Other | Co-Investigator |
| Jovan Tanevski | Other | Co-Investigator |
| Seth Winfree | Other | Co-Investigator |
| Robin Fallegger | Other | Fellow |
| Steve Bogen | Other | Other |
| Charlotte Boys | Other | Other |
| Brandon Ginley | Other | Other |
| Leah Guthrie | Other | Other |
| Leonie Küchenhoff | Other | Other |
| Brendon Lutnick | Other | Other |
| Joseph Ardayfio | Other | Patient Partner,Other |
| Jack Bebiak | Other | Patient Partner,Other |
| Taneisha Campbell | Other | Patient Partner,Other |
| Robert Koewler | Other | Patient Partner,Other |
| Roy Pinkeney | Other | Patient Partner,Other |
| John Saul | Other | Patient Partner,Other |
| Dongwon Lee | Other | Principal Investigator |
| Anant Madabhushi | Other | Principal Investigator |
| Ari Pollack | Other | Principal Investigator |
| Julio Saez-Rodriguez | Other | Principal Investigator |
| Raf Van de Plas | Other | Principal Investigator |
| Ashley R Burg | Other | Research Coordinator,Study staff |
| Lukasz G. Migas | Other | Study staff |

| | | |
|---|---|---|
| Ljiljana Paša-Tolić | Pacific Northwest National Laboratory | Co-Investigator |
| Dusan Velickovic | Pacific Northwest National Laboratory | Co-Investigator |
| Jessica Lukowski | Pacific Northwest National Laboratory | Other |
| Christopher R Anderton | Pacific Northwest National Laboratory | Principal Investigator |
| Kisurb Choe | Pacific Northwest National Laboratory | Study staff |
| Brittney L. Gorman | Pacific Northwest National Laboratory | Study staff |
| George (Holt) Oliver | Parkland Health and Hospital System | Other |
| Rachel S. G. Sealfon | Princeton University | Co-Investigator |
| Xi Chen | Princeton University | Other |
| Weiguang Mao | Princeton University | Other |
| Ksenia Sokolova | Princeton University | Other |
| Aaron Wong | Princeton University | Other |
| Olga G Troyanskaya | Princeton University | Principal Investigator |
| David H. Beyda, MD | University of Arizona | Co-Investigator |
| thajudeen b | University of Arizona | Co-Investigator |
| Rebecca Tsosie | University of Arizona | Co-Investigator |
| Gregory Woodhead | University of Arizona | Co-Investigator,Biopsy Operator |
| Erika R Bracamonte | University of Arizona | Pathologist |
| Raymond Scott | University of Arizona | Patient Partner |
| Frank C. Brosius | University of Arizona | Principal Investigator |
| Baltazar Campos | University of Arizona | Research Coordinator |
| Austin Derma | University of Arizona | Research Coordinator |
| Daniel Damian Duran | University of Arizona | Research Coordinator |
| Griselda Gamez | University of Arizona | Research Coordinator |
| Nicole Marquez | University of Arizona | Research Coordinator |
| Katherine Mendoza | University of Arizona | Research Coordinator |
| Ana Celina Sanora | University of Arizona | Research Coordinator |
| Kun Zhang | University of California San Diego | Co-Investigator |
| Tara K Sigdel | University of California San Francisco | Co-Investigator |
| Kavya Anjani | University of California San Francisco | Other |
| Tariq Mukatash | University of California San Francisco | Other |
| Zoltan G. Laszik | University of California San Francisco | Principal Investigator |
| Minnie M Sarwal | University of California San Francisco | Principal Investigator |
| James G. Cimino | University of California San Francisco | Research Coordinator |
| Dane Munar | University of California San Francisco | Research Coordinator |
| Laura Pyle | University of Colorado | Co-Investigator |
| Julia Wrobel | University of Colorado | Co-Investigator |
| Carissa Vinovskis | University of Colorado | Other |
| Petter Bjornstad | University of Colorado | Principal Investigator |
| Hsieh EWY | University of Colorado | Principal Investigator |
| Joshua M. Thurman | University of Colorado | Principal Investigator |
| Pinaki Sarder | University of Florida | Co-Investigator |
| Manoj Kumar Galla | University of Florida | Other |

| | | |
|---|---|---|
| Harshit Lohaan | University of Florida | Other |
| Sayat Mimar | University of Florida | Other |
| Samuel Border | University of Florida | Study staff |
| Nicholas Lucarelli | University of Florida | Study staff |
| Ahmed Naglah | University of Florida | Study staff |
| Anindya S. Paul | University of Florida | Study staff |
| Michael Tanious | University of Illinois, Chicago | Biopsy Operator |
| Tanika N. Kelly | University of Illinois, Chicago | Co-Investigator |
| Bui, JT or Bui, James T | University of Illinois, Chicago | Co-Investigator,Biopsy Operator |
| Ron C. Gaba | University of Illinois, Chicago | Co-Investigator,Biopsy Operator |
| Aaron Scroggins | University of Illinois, Chicago | Other |
| Suman Setty | University of Illinois, Chicago | Pathologist |
| Monica L. Fox | University of Illinois, Chicago | Patient Partner |
| James P. Lash | University of Illinois, Chicago | Principal Investigator |
| Ana C. Ricardo | University of Illinois, Chicago | Principal Investigator |
| Anand Srivastava | University of Illinois, Chicago | Principal Investigator,Co-Investigator,Other |
| Joed Ancheta | University of Illinois, Chicago | Research Coordinator |
| Eunice Carmona-Powell | University of Illinois, Chicago | Research Coordinator |
| Natalie Meza | University of Illinois, Chicago | Research Coordinator |
| Arabela Quiroga | University of Illinois, Chicago | Research Coordinator |
| Amada Renteria | University of Illinois, Chicago | Research Coordinator |
| Kim Silva | University of Illinois, Chicago | Research Coordinator,Project Manager |
| Devona Redmond | University of Illinois, Chicago | Study staff,Other |
| Fadhl Alakwaa | University of Michigan | Co-Investigator |
| Ulysses G. J. Balis | University of Michigan | Co-Investigator |
| Markus Bitzer | University of Michigan | Co-Investigator |
| Yongqun He | University of Michigan | Co-Investigator |
| Wenjun Ju | University of Michigan | Co-Investigator |
| Laura H. Mariani | University of Michigan | Co-Investigator |
| Rajasree Menon | University of Michigan | Co-Investigator |
| Abhijit S. Naik | University of Michigan | Co-Investigator |
| Edgar A. Otto | University of Michigan | Co-Investigator |
| Jennifer A. Schaub | University of Michigan | Co-Investigator |
| Dawit Demeke | University of Michigan | Fellow |
| Francesca Annese | University of Michigan | Other |
| Heather K. Ascani | University of Michigan | Other |
| Victoria M. Blanc | University of Michigan | Other |
| Nathan Creger | University of Michigan | Other |
| Rachel Dull | University of Michigan | Other |
| Renee Frey | University of Michigan | Other |
| Josh Hartley | University of Michigan | Other |

| | | |
|---|---|---|
| Chrysta C Lienczewski | University of Michigan | Other |
| Becky Steck | University of Michigan | Other |
| Haneen Tout | University of Michigan | Other |
| Zach Wright | University of Michigan | Other |
| Matthias Kretzler | University of Michigan | Principal Investigator |
| Jeffrey B. Hodgin | University of Michigan | Principal Investigator,Co-Investigator,Pathologist |
| Lalita Subramanian | University of Michigan | Project Manager |
| Nikole Bonevich | University of Michigan | Project Manager,Other |
| Ninive Conser | University of Michigan | Study staff |
| Sean Eddy | University of Michigan | Study staff |
| John Hartman | University of Michigan | Study staff |
| Phillip J. McCown | University of Michigan | Study staff |
| Viji Nair | University of Michigan | Study staff |
| Rebecca Reamy | University of Michigan | Study staff |
| Michael P. Rose | University of Michigan | Study staff |
| Cathy Smith | University of Michigan | Study staff |
| Donna D'Souza | University of Minnesota | Biopsy Operator |
| Siobhan M. Flanagan | University of Minnesota | Biopsy Operator |
| Jerica M. Berge | University of Minnesota | Co-Investigator |
| Drawz PE | University of Minnesota | Co-Investigator |
| Tasma Harindhanavudhi | University of Minnesota | Co-Investigator |
| Sisi Ma | University of Minnesota | Co-Investigator |
| Elizabeth A. Rogers | University of Minnesota | Co-Investigator |
| Michael S. Rosenberg | University of Minnesota | Co-Investigator |
| Sami Safadi | University of Minnesota | Co-Investigator |
| Susan M. Wolf | University of Minnesota | Co-Investigator |
| Christopher J. Jones | University of Minnesota | Co-Investigator,Biopsy Operator |
| Sandeep Sharma | University of Minnesota | Co-Investigator,Biopsy Operator |
| Oyedele A. Adeyi | University of Minnesota | Pathologist |
| Yanli Ding | University of Minnesota | Pathologist |
| Ann Gentry | University of Minnesota | Patient Partner |
| Susan Klett | University of Minnesota | Patient Partner |
| M. Luiza Caramori | University of Minnesota | Principal Investigator |
| Patrick H. Nachman | University of Minnesota | Principal Investigator |
| Alyson Coleman | University of Minnesota | Project Manager |
| Dori Henderson | University of Minnesota | Project Manager |
| Cathy A Bagne | University of Minnesota | Research Coordinator |
| Rachel R. Kaspari | University of Minnesota | Research Coordinator |
| Oluwatosin Oluwole | University of Minnesota | Research Coordinator |
| Via Rao | University of Minnesota | Research Coordinator |
| Nicolas J Rauwolf | University of Minnesota | Research Coordinator |
| Michelle L. Snyder | University of Minnesota | Research Coordinator |

| | | |
|---|---|---|
| Zoe Wright | University of Minnesota | Research Coordinator |
| Alison Bunio Alvear | University of Minnesota | Study staff |
| Peter R. Bream, Jr. | University of North Carolina | Biopsy Operator |
| Nicole Keefe | University of North Carolina | Biopsy Operator |
| Priya Mody | University of North Carolina | Biopsy Operator |
| Saad Mohammed Shariff | University of North Carolina | Biopsy Operator |
| Alexander Villalobos | University of North Carolina | Biopsy Operator |
| Prabir Roy-Chaudhury | University of North Carolina | Co-Investigator |
| Evan M. Zeitler | University of North Carolina | Co-Investigator |
| Vanessa Moreno | University of North Carolina | Co-Investigator,Pathologist |
| Samuel Haddad | University of North Carolina | Other |
| J Charles Jennette | University of North Carolina | Pathologist |
| Jennifer L. Jones | University of North Carolina | Patient Partner |
| Amy K. Mottl | University of North Carolina | Principal Investigator |
| Tashas Cameron-Wheeler | University of North Carolina | Research Coordinator |
| Mary M. Collie | University of North Carolina | Research Coordinator |
| Anne Froment | University of North Carolina | Research Coordinator |
| Dhatri Kakarla | University of North Carolina | Research Coordinator |
| Sara S. Kelley | University of North Carolina | Research Coordinator |
| Sora Lee | University of North Carolina | Research Coordinator |
| Fernanda Ochoa Toro | University of North Carolina | Research Coordinator |
| Sandhya Sundar Rajan | University of North Carolina | Research Coordinator,Study staff |
| Matthew Gilliam | University of Pittsburgh | Other |
| Daniel E. Hall | University of Pittsburgh | Other |
| John A. Kellum | University of Pittsburgh | Other |
| Roderick Tan | University of Pittsburgh | Other |
| James Winters | University of Pittsburgh | Other |
| Parmjeet Randhawa | University of Pittsburgh | Pathologist |
| Raghavan Murugan | University of Pittsburgh | Principal Investigator |
| Paul M. Palevsky | University of Pittsburgh | Principal Investigator |
| Matthew R. Rosengart | University of Pittsburgh | Principal Investigator |
| Michele M Elder | University of Pittsburgh | Project Manager |
| Adam Burgess | University of Pittsburgh | Research Coordinator |
| Tina Vita | University of Pittsburgh | Research Coordinator |
| Soumya Maity | University of Texas Health Science Center at San Antonio | Co-Investigator |
| Guanshi Zhang | University of Texas Health Science Center at San Antonio | Co-Investigator |
| manjeri venkatachalam | University of Texas Health Science Center at San Antonio | Co-Investigator,Pathologist |
| Bhupendra Kumar Gurung | University of Texas Health Science Center at San Antonio | Other |

| | | |
|---|---|---|
| Annapurna Pamreddy | University of Texas Health Science Center at San Antonio | Other |
| Hongping Ye | University of Texas Health Science Center at San Antonio | Other |
| Shiqi Zhang | University of Texas Health Science Center at San Antonio | Other |
| Kumar Sharma | University of Texas Health Science Center at San Antonio | Principal Investigator |
| NAGARJUNACHARY RAGI | University of Texas Health Science Center at San Antonio | Study staff |
| Samuel Rice | University of Texas Southwestern | Biopsy Operator |
| S. Susan Hedayati | University of Texas Southwestern | Co-Investigator |
| Meredith C McAdams | University of Texas Southwestern | Co-Investigator |
| R. Tyler Miller | University of Texas Southwestern | Co-Investigator |
| Jiten Patel | University of Texas Southwestern | Co-Investigator |
| Choudhary Moaz | University of Texas Southwestern | Co-Investigator,Biopsy Operator |
| Sanjeeva P. Kalva | University of Texas Southwestern | Co-Investigator,Biopsy Operator |
| Allen R Hendricks | University of Texas Southwestern | Co-Investigator,Pathologist |
| Asra Kermani MD | University of Texas Southwestern | Other |
| Simon C. Lee | University of Texas Southwestern | Other |
| Harold Park | University of Texas Southwestern | Other |
| Anil Pillai | University of Texas Southwestern | Other |
| Natasha Wen | University of Texas Southwestern | Other |
| Qi Cai | University of Texas Southwestern | Pathologist |
| Jose R. Torrealba | University of Texas Southwestern | Pathologist |
| Catherine Campbell | University of Texas Southwestern | Patient Partner |
| Robert D Toto | University of Texas Southwestern | Principal Investigator |
| Miguel A. Vazquez | University of Texas Southwestern | Principal Investigator |
| Shihong Ma | University of Texas Southwestern | Research Coordinator |
| Nancy Wang | University of Texas Southwestern | Research Coordinator |
| Boris S. Patlis | University of Texas Southwestern | Research Coordinator,Other |
| Andrew N Hoofnagle | University of Washington | Co-Investigator |
| Robyn L. McClelland | University of Washington | Co-Investigator |
| Kasra A Rezaei | University of Washington | Co-Investigator |
| Jaime Snyder | University of Washington | Co-Investigator |
| Katherine R. Tuttle | University of Washington | Co-Investigator |
| Bessie A. Young | University of Washington | Co-Investigator |
| Christine P Limonte | University of Washington | Co-Investigator,Fellow |
| Ruikang Wang | University of Washington | Co-Investigator,Other |
| Stephanie M. Grewenow | University of Washington | Other |
| Cienn N. Joyeux | University of Washington | Other |
| Yunbi Nam | University of Washington | Other |
| Christopher Park | University of Washington | Other |

| | | |
|---|---|---|
| Adam Wilcox | University of Washington | Other |
| Kayleen Williams | University of Washington | Other |
| CE Alpers | University of Washington | Pathologist |
| Kelly D. Smith | University of Washington | Pathologist |
| Keith D. Brown | University of Washington | Patient Partner |
| Lynda Hayashi | University of Washington | Patient Partner |
| Nichole M. Jefferson | University of Washington | Patient Partner |
| Richard A. Knight | University of Washington | Patient Partner |
| Glenda V. Roberts | University of Washington | Patient Partner |
| Christy Stutzke | University of Washington | Patient Partner |
| Ian H. de Boer | University of Washington | Principal Investigator,Co-Investigator |
| Brooke Berry | University of Washington | Project Manager |
| Kristina N Blank | University of Washington | Project Manager |
| Ashveena L Dighe | University of Washington | Project Manager |
| Alexa Plisiewicz | University of Washington | Project Manager |
| Natalya Sarkisova | University of Washington | Project Manager |
| Ashley C Berglund | University of Washington | Study staff |
| Jonas M Carson | University of Washington | Study staff |
| Matthew Dekker | University of Washington | Study staff |
| Frederick Dowd | University of Washington | Study staff |
| Jimmy Phuong | University of Washington | Study staff |
| Artit Wangperawong | University of Washington | Study staff |
| de Caestecker M.P. | Vanderbilt University | Co-Investigator |
| Agnes B. Fogo | Vanderbilt University | Co-Investigator,Pathologist |
| Katerina V. Djambazova | Vanderbilt University | Fellow |
| Audra M. Judd | Vanderbilt University | Other |
| Yarieli Cuevas-Rios | Vanderbilt University | Patient Partner |
| Yuankai Huo | Vanderbilt University | Principal Investigator |
| Jeffrey M. Spraggins | Vanderbilt University | Principal Investigator |
| Melissa A Farrow | Vanderbilt University | Project Manager |
| Jamie L Allen | Vanderbilt University | Study staff |
| Madeline E. Colley | Vanderbilt University | Study staff |
| Ruining Deng | Vanderbilt University | Study staff |
| Martin Dufresne | Vanderbilt University | Study staff |
| Angela R.S. Kruse | Vanderbilt University | Study staff |
| Jeannine Basta | Washington University St. Louis | Co-Investigator |
| Anitha Vijayan | Washington University St. Louis | Co-Investigator |
| Reetika Ghag | Washington University St. Louis | Other |
| Amanda Knoten | Washington University St. Louis | Other |
| Asmita L | Washington University St. Louis | Other |
| Stephanie Reinert | Washington University St. Louis | Other |
| Joseph P. Gaut | Washington University St. Louis | Pathologist |

| | | |
|---|---|---|
| Michael Rauchman | Washington University St. Louis | Principal Investigator |
| Sanjay Jain | Washington University St. Louis | Principal Investigator,Other |
| Madhurima Kaushal | Washington University St. Louis | Project Manager,Study staff |
| Amy McMurray | Washington University St. Louis | Research Coordinator |
| Kristine Conlon | Washington University St. Louis | Research Coordinator,Patient Partner |
| Brittany C Minor | Washington University St. Louis | Study staff |
| Gerald Nwanne | Washington University St. Louis | Study staff |
| Bo Zhang | Washington University St. Louis | Study staff |
| Jeffrey M Turner | Yale University | Biopsy Operator |
| Moledina DG | Yale University | Co-Investigator |
| Tanima Arora | Yale University | Other |
| Gilbert W. Moeckel | Yale University | Pathologist |
| Vijayakumar R Kakade | Yale University | Principal Investigator |
| Lloyd G Cantley | Yale University | Principal Investigator,Co-Investigator |
| F. Perry Wilson | Yale University | Principal Investigator,Co-Investigator |
| Ugochukwu Ugwuowo | Yale University | Research Coordinator |
| Angela M. Victoria-Castro | Yale University | Research Coordinator |
| Melissa M. Shaw | Yale University | Research Coordinator,Study staff |
| Tifanny Budiman | Yale University | Study staff |
